# CausalCellInfer: Resolving cell-type-specific disease mechanisms from biobank-scale GWAS

**DOI:** 10.1101/2024.10.17.24315646

**Authors:** Liangying Yin, Yujia Shi, Ruoyu Zhang, Yong Xiang, Jinghong Qiu, Pak-Chung Sham, Hon-Cheong So

## Abstract

Integrating the cellular resolution of single-cell RNA sequencing (scRNA-seq) with the phenotypic depth of population-scale biobanks is essential for elucidating the cellular basis of complex diseases. However, this integration is often hindered by the limited sample sizes of scRNA-seq cohorts and the lack of cell-type resolution in massive biobank datasets.

We present CausalCellInfer, a scalable computational framework designed to bring cellular resolution to bulk and genotype-imputed transcriptomes. CausalCellInfer utilizes an invariant causal prediction-inspired procedure (scI-GCM) to identify environment-stable marker genes, employs a parsimonious deep neural network for robust cell-fraction deconvolution, and leverages regularized matrix completion to reconstruct cell-type-specific (CTS) expression profiles. This architecture is specifically optimized for biobank-scale data, where technical heterogeneity and limited gene overlap are prevalent.

Validated across simulated data, pseudo-bulk mixtures, and real PBMC datasets, CausalCellInfer demonstrated superior accuracy and computational efficiency compared to existing methods. Applied to ∼500,000 UK Biobank participants, the framework enabled cell-resolved analyses for 29 traits, identifying known pathological shifts, such as reduced pancreatic β-cell proportions in Type 2 Diabetes, and uncovering novel biological signals, including disrupted excitatory neuron and oligodendrocyte interactions in depression. Furthermore, inferred CTS differential expression patterns showed significant concordance with independent single-cell studies and were enriched for OpenTargets disease genes. Overall, CausalCellInfer bridges the gap between single-cell insights and population-scale genomics, providing a powerful tool for systematic discovery of disease mechanisms at cellular resolution.

## Introduction

Single-cell RNA-sequencing (scRNA-seq) has transformed our understanding of cellular diversity and disease biology by enabling gene expression profiling at single-cell resolution^1–4^. However, routine application of scRNA-seq in large cohorts remains limited by cost, technical complexity, and often sparse clinical data on symptoms, prognosis, and comorbidities. Most single-cell studies are designed for a specific disease or question, and therefore often include limited subject-level clinical information (e.g., symptoms, prognosis, comorbidities, medications) and small numbers of individuals. These constraints make it difficult to connect cellular mechanisms to diverse clinical outcomes, to systematically assess effect modifiers, or to distinguish causes from consequences of disease.

In contrast, bulk RNA-seq and genotype-imputed transcriptome data are available at population scale. Resources such as TCGA and GTEx offer bulk expression across tissues, and large biobanks like the UK Biobank (UKBB) provide extensive genotype data together with deep clinical and biological phenotypes. Genotype data further enable the prediction of individual-level gene expression, creating transcriptome-like measurements at biobank scale^5–7^. However, both measured bulk expression and genetically predicted expression lack cellular resolution. Bulk expression profiles aggregate signals across heterogeneous cell types, obscuring the cell-type-specific (CTS) drivers of complex traits. Bridging the mechanistic resolution of scRNA-seq with the statistical power and phenotypic breadth of biobanks would enable cell-type–resolved discovery in large cohorts and across a variety of clinical outcomes. We address this gap with **CausalCellInfer**, a framework inspired by the concept of invariant causal prediction (ICP), as proposed by Peters et al.^8^. CausalCellInfer uses scRNA-seq reference data to identify robust, cell-defining features and then transfers this information to bulk transcriptomes, including genotype-imputed expression in large cohorts, to infer *(i) cell-type proportions and (ii) CTS expression*.

The design goal is biobank readiness: the proposed framework is scalable to very large sample sizes and reliable under distribution shifts. By anchoring inference at the cell-type level, CausalCellInfer enables biobank-scale analyses of how cellular composition and CTS expression relate to risk factors, disease onset / progression, prognosis, and treatment response. Although the framework can be used with any bulk transcriptome data, one of its most powerful applications arise in biobanks where broad phenotyping, large sample size, and individual-level covariates enable rigorous, cell-type-resolved association analyses.

Using inferred cell-type proportions and CTS expression, we support an application suite that is difficult to implement with conventional single-cell study designs because of limited subject-level phenotyping and small sample sizes:

- **Cell-type specific TWAS (CTS-TWAS)**: We test whether genetically predicted CTS expression is associated with complex traits/diseases, localizing gene–trait associations to the most relevant cell types.
- **CTS-TWAS with causal gene prioritization (CTS-TWAS-Causal):** By integrating causal feature selection, we prioritize genes whose CTS expression is most consistent with a direct effect on traits, helping to distinguish core drivers from correlated signals.
- **Cell-type proportion association studies:** We assess whether genetically inferred cell-type abundances relate to disease risk or clinical phenotypes. This highlights cellular contributors to pathophysiology (e.g., expansion or depletion of specific immune or stromal populations) and nominates candidate cell types for therapeutic targeting.
- **Cell-type proportion interaction analyses**: We evaluate interactions within and across tissues (e.g., immune–parenchymal crosstalk) to identify synergistic or antagonistic relationships that affect disease expression, prognosis, or treatment response.
- **Conditional TWAS:** We adjust bulk or imputed expression analyses for inferred cell-type proportions to reduce confounding by cellular composition when identifying differentially expressed genes. This improves interpretability by separating compositional shifts from CTS expression changes.

These analyses operate on individual-level genotype and imputed expression, and also scale readily to thousands of participants and thousands of phenotypes, typical in biobanks. This scale yields several practical advantages. First, comprehensive covariates available in biobanks (e.g., age, sex, BMI, smoking, medications, socioeconomic factors) can be included to reduce confounding and to test effect modification. Second, large samples naturally support interaction models, stratified analyses, and prediction frameworks that are underpowered in typical single-cell cohorts. Third, when expression and cell-type measures are genetically predicted (from germline variations), associations are less susceptible to reverse causation and time-varying confounders (e.g., disease severity, treatment), strengthening causal interpretation relative to observational single-cell measurements. While not a definitive proof of causality, this approach helps infer whether cellular features are more likely antecedents rather than consequences of a disease.

Applying CausalCellInfer to UKBB dataset, we connect cell-type proportions and CTS expression to a wide spectrum of clinical phenotypes, from cardiometabolic traits and immune disorders to neurological diseases. This biobank-scale setting enables us to: (i) map cell types most strongly linked to specific outcomes; (ii) nominate CTS genes whose dysregulation associates with disease liability; and (iii) reveal cellular interactions that may underlie comorbidities or variable treatment response. We further assess biological plausibility by demonstrating significant overlaps between CTS differentially expressed genes inferred from UKBB and those reported in independent scRNA-seq studies, providing external validation across diseases and tissues.

Here we highlight key differences and strengths of our proposed approach relative to several existing deconvolution methods. Signature-based deconvolution methods (e.g. CIBERSORTx, DWLS) rely on reference gene expression profiles for marker genes derived from scRNA-seq^9–14^. However, applying signature-based approaches to genotype-imputed transcriptomes is often challenging because reliable imputed expression is available for only a *subset* of genes (typically on the order of a few thousand, depending on tissue and prediction models), which can remove or weaken key markers and reduce identifiability of cell types. Also, while signature-based methods can perform well when the reference and target data are well matched, in practice their accuracy can degrade under technical and biological mismatches (e.g., batch effects, platform differences, disease states, subject heterogeneity etc.) because the marker set and reference profiles may not transfer very well across settings.

Signature-free, deep learning–based methods (e.g., Scaden, TAPE) improve robustness by learning directly from single-cell-derived training data^15–17^, but they typically require high-dimensional features as input and substantial computational resources. Also, their performance may be sensitive to how well the training distribution matches the target cohort, creating risks of overfitting or reduced generalization under distribution shifts^18–21^.

Our framework is designed for the biobank use case: it focuses on identifying a parsimonious set of cell-defining features that are stable across environments (motivated by invariance ideas^8,22^), and it explicitly targets transfer to bulk and genotype-imputed expression where feature availability may be constrained. By prioritizing transferable, compact feature sets and emphasizing out-of-cohort reliability, our framework is intended to remain accurate across cohorts with different characteristics (e.g. demographics, technologies, disease burdens) while scaling to very large samples and thousands of phenotypes. These design choices are driven not by methodological novelty per se, but by the practical need to execute cell-type–resolved analyses across very large samples and many phenotypes.

In summary, the major objective of CausalCellInfer is to bring cellular resolution to biobank-scale discovery. By inferring cell-type proportions and CTS expression from bulk and genotype-imputed transcriptomes, we enable:

- Systematic **CTS-TWAS** to localize gene–trait associations to the most relevant cell types;
- Prioritization of putatively **causal CTS genes** for mechanistic studies and therapeutic target nomination;
- Association and interaction analyses of **cell-type abundances** within and across tissues; and
- Confounding-aware differential expression through **conditional TWAS**.

Together, these capabilities allow investigators to interrogate disease mechanisms at the cellular level while retaining the statistical power, phenotypic breadth, and covariate control afforded by large cohorts.

## Methods

### Overview of CausalCellInfer

In this study, we introduce an integrated framework, CausalCellInfer, that combines invariant causal prediction and DNNs with scRNA data to estimate cell proportions and expression profiles from bulk or genotype-imputed expression data across large cohorts.

Given (1) a reference scRNA-seq dataset with cell-type labels and (optionally) ‘environment’ annotations and (2) a target cohort with bulk or genetically-predicted expression, CausalCellInfer produces two outputs for each target individual: **estimated cell-type proportions** and **cell-type-specific (CTS) expression profiles.**

The proposed workflow consists of three main steps (Fig. 1):

1. **Invariant cell-marker identification** from the scRNA-seq reference using an invariant causal prediction (ICP)-inspired feature selection procedure (scI-GCM).
2. **Cell-type deconvolution** in the target cohort using a deep neural network (DNN) trained on pseudo-bulk mixtures generated from the scRNA-seq reference, restricted to the selected marker genes.
3. **CTS expression reconstruction** using a regularized matrix completion approach (ENIGMA) informed by the estimated cell proportions and scRNA-seq reference profiles.

**Figure 1.**
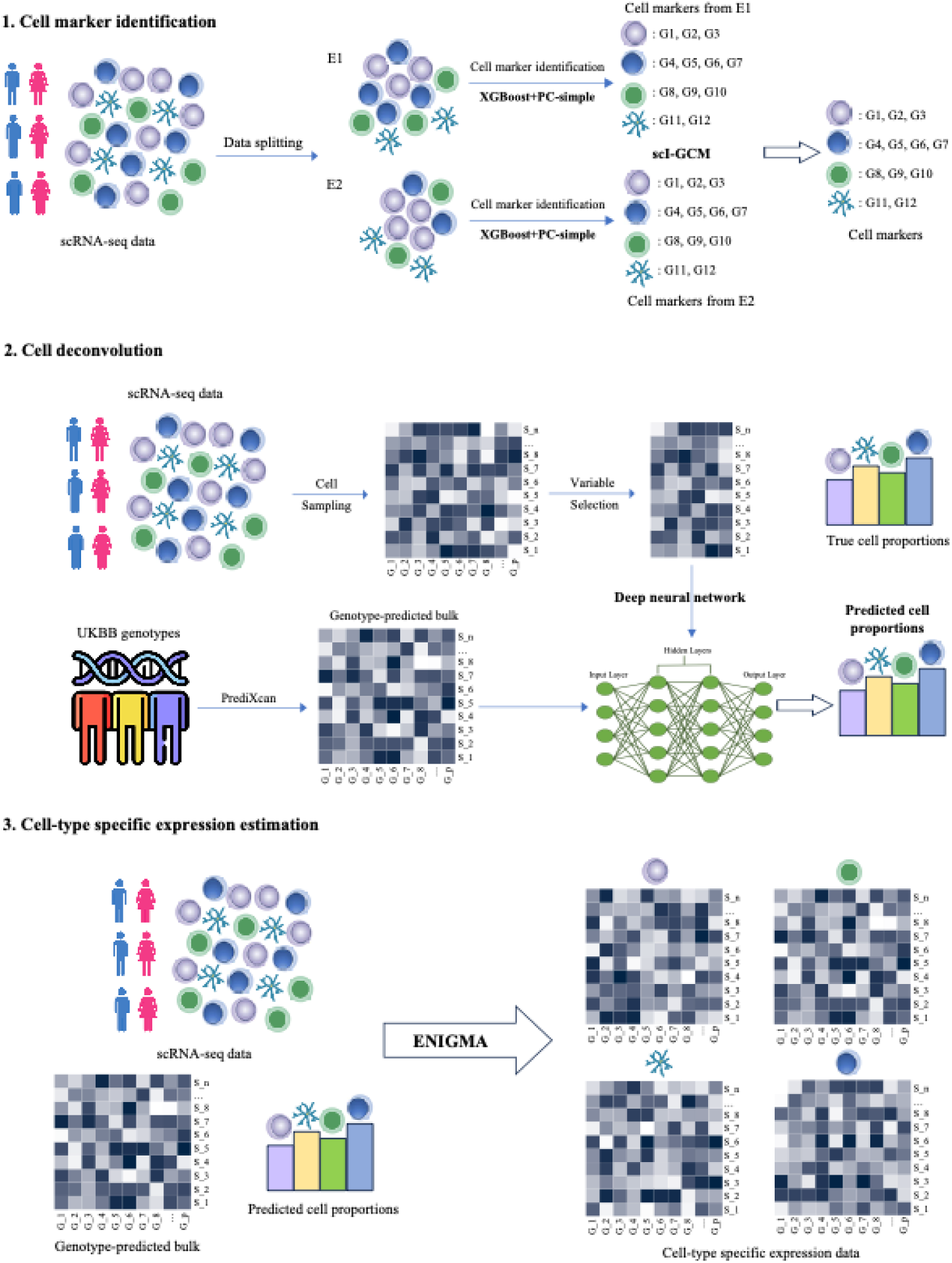
Workflow of our proposed framework for cell decomposition and cell type-specific expression estimation. 1. **Cell marker identification**: We proposed invariant generalized covariance measure (scI-GCM) to identify cell markers for each cell type from the reference scRNA-seq data; 2. **Cell deconvolution**: We modified Scaden, a DNN-based deconvolutional method, to estimate cell proportions from bulk expression data using a small yet informative gene set; 3. **Cell type-specific expression estimation**: We employed a regularized matrix completion method (ENIGMA) to reconstruct cell type-specific expression data by leveraging information from the reference RNA-seq data and the estimated cell proportions from step 2.

Subsequent sections will provide a detailed description of each step.

### Step 1 - Cell marker identification

Genes used as “markers” in scRNA-seq are usually cell-type enriched, but they can still vary substantially with disease states, age, sex, technical batch, or other sample-specific factors. For deconvolution and transfer to external cohorts, we seek a *parsimonious* set of cell-defining genes whose relationship to cell identity is stable across environments.

Robust cell markers tend to exhibit stability across diverse environments. In this context, we define “environments” (*E*) as subject-related or experimental background features that may shift expression distributions. These environments may include variations in disease pathology, demographics, spatiotemporal factors, and batch effects, alongside experimental perturbations like gene knockdown or overexpression. However, the “environment” should not include the outcome variable and its descendants^8^. According to the theory of Invariant Causal Prediction (ICP), the conditional distribution of a target variable *Y* is invariant across environments when conditioned on its direct causes.

Operationally, we seek to identify a gene set 𝑋^𝑆^ such that the association between cell-type 𝑌 and 𝐸 disappears after conditioning on 𝑋^𝑆^, satisfying the invariance criterion:

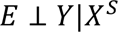

Under standard ICP assumptions, this independence ensures that 𝑋^𝑆^ captures the fundamental, stable signals defining cell identity.

To identify such markers within large-scale scRNA-seq data, we introduce an *adapted* and improved Invariant Generalized Covariance Measure (I-GCM) framework for **s**ingle-**c**ell analysis, which we also refer to as “scI-GCM” below. The presented approach refines our previously proposed I-GCM framework^23^, originally developed to identify clinical and genetic risk factors in epidemiological cohorts^23^. Here we optimized the I-GCM pipeline to tackle the high-dimensional challenges inherent in single-cell transcriptomic data. The resulting framework is grounded in causal graphical models and invariant prediction principles, operating under standard causal assumptions such as partial faithfulness, unconfoundedness, and stable causal mechanisms. Crucially, the method incorporates a scalable, hierarchical feature selection process to ensure computational efficiency.

Below we summarize the key steps and then describe adaptations specific to cell marker selection:

1. **Candidate marker identification via two-stage hierarchical screening** To address the computational intensity of standard causal discovery, we implemented a two-stage hierarchical screening process:

i. ***Initial Screening (XGBoost)***: The scRNA-seq data was first partitioned into training and testing sets. The training set was then subdivided into subsets based on selected environmental variables, specifically disease status and gender for this study. Within each environment stratum, we trained an XGBoost model (max tree depth 2, learning rate 1) to predict the binary cell type (𝑌) from gene expression data (𝑋). This step performs a “coarse” reduction, retaining genes with positive importance scores.
ii. ***Causal Refinement (PC-simple)***: To eliminate spurious associations captured by XGBoost, we refine the retained genes using the PC-simple algorithm within each environment stratum. We set the maximum conditioning order to 3 and significant level (𝛼) to 0.05. The union of genes across all environments formed the initial candidate set to be analyzed in the next stage. Regarding the principles of PC-simple, it is a causal inference algorithm that generalizes ordered correlation screening to select direct causal variables. A candidate gene 𝑋^𝑗^ is retained if the null hypothesis of conditional independence between 𝑌 and 𝑋^𝑗^ given a conditioning set 𝑋^𝑆^ is rejected using Fisher’s Z-transformed partial correlation:

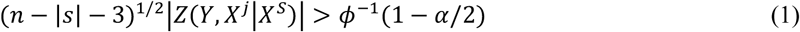

where 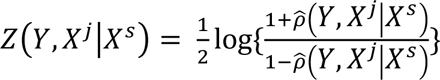 denotes Fisher’s Z-transform of the partial correlation, 𝑛 denotes the sample size, |𝑠| is the conditioning set size, 𝜙^−1^indicates the is the standard normal quantile function.
2. **Invariant marker identification via generalized covariance measure (GCM)** The final stage identifies markers that define cell identity amidst environmental noise. We employed the generalized covariance measure (GCM) (𝑇^𝑛^) ^24^ to test whether the conditional association between the environment variable(𝐸) and cell type(𝑌) remains stable when conditioning on a specific gene set (𝐸 ⊥ 𝑌|𝑋^𝑆^). The GCM statistic(𝑇^𝑛^) tests independence based on residuals(𝑅):

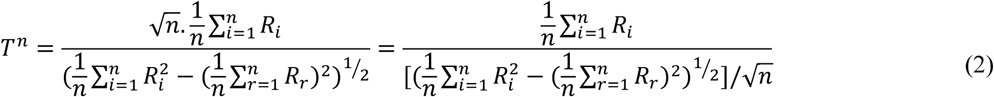

Here 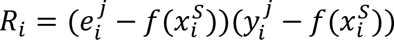 represents the product between residuals from the prediction functions of 𝐸 and Y using 𝑋^𝑆^ for each observation. We utilize XGBoost as the prediction model to calculate this conditional independence; genes are iteratively removed if their inclusion does not contribute to improving They would have gone the invariance. Other machine learning methods can also be used as substitute for XGBoost, offering high flexibility. Significant deviation from the null hypothesis(indicating a change in GCM distance between consecutive conditional gene sets) is rejected if:

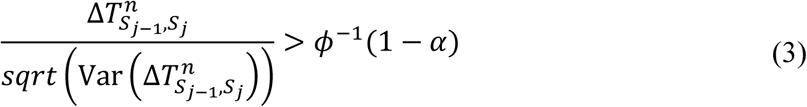

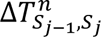 defines the change in the distance of GCM from zero between two consecutive conditional sets, with 𝛼 = 0.05. For more technical details, please refer to supplementary text.

### Step 2: Cell-type deconvolution with ensemble DNN

#### Training data generation (pseudo-bulk mixtures)

In our framework, we employ a deep neural network (DNN) model to estimate cell proportions. It uses simulated bulk gene counts for the selected marker genes derived from reference scRNA-seq data to estimate cell proportions in bulk expression samples.

We train a DNN to map bulk expression of the selected marker genes to cell-type fractions. Because the target cohort may be bulk RNA-seq or genotype-imputed expression, we train using pseudo-bulk mixtures generated from the scRNA-seq reference, following established pseudo-bulk simulation procedures.

To train the DNN models, we simulated 5,000 pseudo-bulk samples with random fractions of different cell types (𝑓_𝑐_ 𝜖 [0,1] 𝑓𝑜𝑟 𝑎𝑙𝑙 𝑐 𝜖{1,2, … 𝐶}, 𝑤ℎ𝑒𝑟𝑒 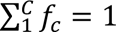) by subsampling cells from the reference RNA-seq data, following the pseudo-bulk data generation approach by Chen et al.^16^. Only genes selected by our proposed scI-GCM approach were included as model features. Each simulated bulk sample was constructed from 500 randomly sampled cells. The number of cells for each cell type in each bulk sample was 𝑟_𝑐_ = 𝑓_𝑐_ ⋅ 500, where 𝑟_𝑐_ refers to the cell number of cell type 𝑐 and 𝑓_𝑐_ refers to the fraction of cell type 𝑐. We generated the final pseudo-bulk expression profile by summing up the single-cell expression levels for each sample.

### Model architecture and training

We used the MinMaxScaler() class from Sklearn to scale the input data to the range [0,1]. We trained an ensemble of 3 DNN models with distinct architectures and degrees of dropout regularization, as the Ensemble DNN model has been shown to outperform individual models^15^. Each model outputs a 𝐶-dimensional fraction vector via a Softmax output layer.

We optimize the mean absolute error (L1 loss):

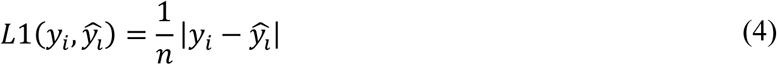

where 𝑦_𝑖_ is the vector of ground-truth cell fractions of the pseudo-bulk sample 𝑖, 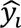 is the vector of predicted fractions of the sample 𝑖, and *n* is the sample size. The final estimated proportions are the ensemble average:

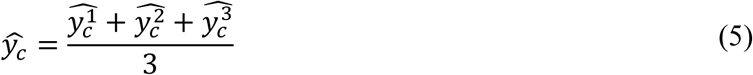

Models were implemented in PyTorch. Trained models are then applied to target bulk (or calibrated imputed) expression to estimate cell-type proportions per individual.

### Step 3: CTS expression estimation

We employed a regularized matrix completion-based approach called ENIGMA^25^ to estimate cell-type specific expression profiles based on predicted cell proportions and genotype-imputed bulk expression profiles. ENIGMA is developed based on two assumptions. The first assumption is that the expectation of each cell type-specific (CTS) expression profile aligns with the reference profile matrix from the external scRNA-seq data^26^. The second assumption is that observation noise is consistent across different samples. We estimate the CTS expression by the trace norm model, following^25^:

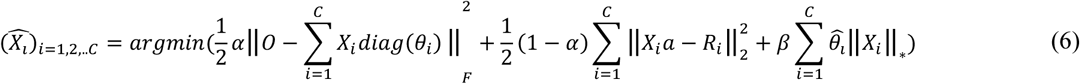

where

𝑖 denotes the cell type index,
𝑂 ∈ 𝑅^𝑚×𝑛^ denotes the bulk count expression matrix for 𝑚 genes and 𝑛 subjects,
𝑋_𝑖_ denotes the true CTS expression for the 𝑖th cell type,
𝜃_𝑖_ denotes the cell proportion of the 𝑖th cell type,
𝑅_𝑖_ denotes the reference expression vector for the 𝑖th cell type,
𝛼 𝜖(0,1) denotes the weights of the aggregated CTS expression profiles (note that 1 − 𝛼 represents the ‘agreement weight’ between the aggregated CTS expression profiles and its corresponding reference expression vector),
𝛽 ∈ (0, +∞) denotes the regularization strength,
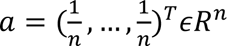 with 𝑛 indicating bulk sample size.

To reduce the computational complexity of the optimization problem, we employed the maximum 𝐿_2_ norm model to deconvolute bulk samples ^27^:

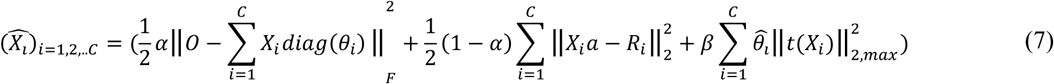

where 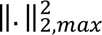 indicates the square of the maximum 𝐿_2_ row norm of a matrix. The optimization procedure was described in^27^.

### Calibrating PrediXcan-imputed expression

PrediXcan-imputed expression values are typically standardized per gene across individuals (often reported as Z-like scores). This per-gene standardization preserves between-individual variation for a given gene, but obscures cross-gene magnitudes *within* an individual. In other words, PrediXcan outputs are suitable for comparing a single gene across individuals (e.g., a value of 1 for a gene in person B indicates higher expression than a value of 0 for the same gene in person A), but they do not reflect actual expression differences between genes within the same individual (e.g., a value of 0 for gene A and −0.3 for gene B in the same person cannot be directly compared). Consequently, the scale of the PrediXcan data is incompatible with scRNA-seq reference data, preventing direct inference of cell composition and cell-type-specific expression.

To mitigate this limitation, we mapped the PrediXcan-imputed values back into the expression space of the scRNA-seq reference, using a heuristic pseudo-bulk calibration. Specifically, we generated 1,000 pseudo-bulk RNA-seq samples from the control-group scRNA-seq data using a procedure similar to that used in the TAPE approach^16^:

1. We took the reference cell-type proportions as a baseline. For each pseudo-bulk sample, we perturbed these proportions by adding Gaussian noise to each cell type, clipped negatives to zero, and renormalized to sum to one.
2. We then sampled 1,000 cells from the scRNA-seq reference with probabilities given by the perturbed proportions and summed their profiles to form one pseudo-bulk sample.

From these 1,000 pseudo-bulk samples, we computed, for each gene 𝑔, the mean 𝜇_𝑔_ and standard deviation 𝜎_𝑔_. For each subject *s*, let *Z_g,s_* denote the PrediXcan-imputed standardized value for gene *g.* We define the calibrated pseudo-bulk-like value:

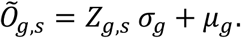

We use 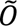 as the bulk input 𝑂 for both DNN deconvolution and ENIGMA reconstruction.

After obtaining the re-scaled bulk RNA-seq data (𝑂), we can derive the reference profile matrix 𝑅 by averaging the expression profiles of cells within each cell type in the scRNA-seq reference. Subsequently, the inferred cell type proportion matrix (𝜃), reference profile matrix (𝑅), and the re-scaled bulk RNA-seq data (𝑂) were plugged into the above formula to infer the final CTS expression profile. The quality of the inferred CTS expression profiles can be significantly influenced by the reference CTS expression profiles. To ensure the reliability of these estimated profiles for subsequent analyses, we recommend using those inferred from the cell types in the reference scRNA-seq dataset with a total cell count exceeding 2,000. Additionally, we aimed to balance the reconstructed CTS so that it resembles the reference scRNA-seq dataset, while ensuring the reconstructed bulk RNA-seq data approximates the observed bulk data. Therefore, we set α to 0.5 in our analysis and kept β at the default setting.

### Evaluation of the proposed framework

#### Simulations to evaluate scI-GCM for single-cell analysis

To validate the advantages of the scI-GCM method, we first established a baseline simulation using simulated scRNA-seq datasets confounded by environmental variables. We compared scI-GCM’s performance against analogous algorithms (PC-simple) and state-of-the-art deconvolution methods (DWLS, Scaden). Notably, the analogous PC-simple method refers to our approach but only replacing scI-GCM in step 1 with PC-simple. Our objective was to test whether scI-GCM more accurately identifies cell-type-specific gene markers in the presence of confounding factors, thereby leading to more robust predictions of cell type proportions.

We utilized scDesign3^27^ to create scRNA-seq data based on a publicly available peripheral blood mononuclear cells (PBMC) dataset. scDesign3 is a statistical simulator that generates realistic single-cell data using a unified probabilistic model, the parameters of which are derived from actual data. This model allows us to manipulate interpretable parameters, introducing environmental effects that lead to confounding, and subsequently simulating new scRNA-seq and bulk data impacted by these effects. We conducted simulations on bulk samples comprising four cell types with varying confounding effects. For more details, please refer to the supplementary text.

#### Evaluation of the proposed approach on real and pseudo-bulk data

To verify the reliability of our proposed framework, we compared the performance of our method with several existing deconvolution approaches across various evaluation metrics, including root mean square error (RMSE), mean absolute error (MAE), and Lin’s Concordance Correlation Coefficient (CCC).

We used pseudo-bulk expression profiles generated from reference scRNA-seq data of four different tissues: the frontal cortex, pancreas islet, and subcutaneous and visceral adipose tissues. These pseudo-bulk expression profiles were simulated in silico from scRNA-seq data with randomly generated cell type proportions, essentially a summation of single-cell expression profiles^16^. We added noise to each generated pseudo-bulk expression profile to mimic the measurement errors in real RNA-seq data^16^. The noise was randomly generated from a Gaussian distribution based on the cells used to create the corresponding pseudo-bulk profile. We only included genes that could be reliably predicted from genotypes by PrediXcan for model training.

Additionally, we conducted benchmarking on real datasets, using available bulk PBMC RNA-seq data and their corresponding cell proportions assessed by flow cytometry from Monaco et al.^28^. To generate pseudo-bulk datasets for data training and to perform scI-GCM to identify cell markers, we merged five PBMC scRNA-seq datasets obtained from different individuals, and these datasets were all downloaded from the 10x Genomics database.

To mitigate overfitting, we partitioned each reference scRNA-seq dataset into two subsets based on subject ID: a training set and a testing set. We initially trained our deep neural network model using the training set and subsequently tested it with pseudo-bulk samples generated from the testing set. The training set was also used to obtain references for MuSiC^29^, DWLS^30^, and Scaden^15^.

#### Validating estimated CTS expression profiles via differential expression analysis

To further validate the robustness of our proposed framework, we performed a comparative analysis on its ability to detect differentially expressed genes across different cell types, especially when cell-type specific (CTS) expression profiles were imputed from genotypes.

Specifically, we identified differentially expressed (DE) genes from both actual scRNA-seq data and genotype-predicted CTS expression profiles. For raw scRNA-seq data, we utilized the FindMarkers() function from the Seurat^31^ package to detect DE genes using the default settings. This function by default employs the Wilcoxon Rank Sum test to identify genes differentially expressed between two cell groups. For our genotype-predicted CTS expression profiles, we applied the same test to detect DE genes. We used positive predictive value (PPV) for the evaluation, considering the DE genes (p-value<=0.05) identified from the actual scRNA-seq dataset as the reference set.

### UK Biobank analyses

#### Cell-type proportion association and interaction analyses with clinical phenotypes

We tested associations between inferred cell-type proportions and disease status/phenotypes using regression models, adjusting for relevant covariates and controlling false discovery rate (FDR) at 0.05 via the Benjamini–Hochberg procedure. Besides, we examined whether interactions between cell types, including cell types *across* different tissues, could affect disease phenotypes using regression models.

#### Identifying cell type-specific genes involved in disease susceptibility (CTS-TWAS) and causal gene prioritization

Upon acquiring the CTS expression profiles for each target disease/trait from UKBB, we identified genes contributing to disease susceptibility for each cell type. This may be considered as a cell type-specific TWAS (CTS-TWAS). To mitigate potential confounding effects from population stratification, we adjusted the estimated cell type-specific expression profiles and phenotypes using the top 10 principal components (PCs) derived from the corresponding genotype dataset.

Subsequently, we utilized the PC-simple algorithm to identify potential direct causal genes for the outcome variable. For binary and count data, we applied PC-simple using linear regression-based partial correlations as an approximation, treating the outcome as a continuous variable. This follows common practice in high-dimensional causal discovery^5,32^. Following ref^32^, we considered an alpha threshold of 0.05 and an order of 3 for the correlation tests. For a more comprehensive description of the algorithm, please refer to ref^5,32^ and the supplementary text.

#### Uncovering cell type-specific genes involved in disease prognosis

UKBB is a comprehensive database that provides extensive phenotypic and prognostic information on various diseases. With the CTS profiles, we could probe the involvement of cell-specific genes in disease *prognoses*, such as mortality and hospitalization frequency. Our application to UKBB mainly focused on 4 diseases/outcomes with imputed expression in relevant tissues (i.e., depression in frontal cortex, T2DM in pancreas, and obesity in subcutaneous and visceral adipose).

We concentrated on key prognostic variables of clinical significance for these diseases. Specifically, we focused on treatment resistance and psychiatric hospitalization frequency for depression (tissue studied: frontal cortex); mortality and related hospitalization frequency for T2DM (tissue studied: pancreas); coronary artery disease (CAD) mortality and related hospitalization frequency for obesity (tissue studied: subcutaneous and visceral adipose). Please refer to the supplementary text for more details.

Moreover, we conducted analyses for 12 other related disorders/phenotypes, including psychosis, bipolar disorder, anxiety (tissue studied: frontal cortex); type 1 diabetes mellitus (T1DM), prediabetes, T2DM-related hospitalization, T2DM requiring insulin (tissue studied: pancreas); CAD, T2DM, heart failure, hypertension, stroke (tissue studied: subcutaneous and visceral adipose). We adjusted the CTS profiles and target prognosis variables by the top 10 PCs to mitigate population stratification. We then employed PC-simple to identify potentially direct causal genes contributing to disease outcomes.

#### Enrichment analysis

We further validated our methodology by assessing whether the identified gene set was significantly enriched among disease genes listed in an independent, established database. We leveraged the OpenTargets database^33^, which provides an overall association score for each gene, aggregating seven types of evidence: genetic associations, somatic mutations, known drugs, affected pathways, RNA expression, literature support, and animal models. Enrichment was performed at the disease level by aggregating CTS genes across cell types.

For further functional interpretation of prioritized (putatively causal) genes, we performed pathway over-representation analysis using ConsensusPathDB^34^ with hypergeometric testing.

#### Conditional TWAS controlling for cell-type proportions

We first performed a standard TWAS analysis on the tissue-specific expression data for each target trait. Then, we examined whether controlling for the estimated cell proportions would significantly affect the effect size of trait-associated genes. Cell proportion is a potential confounder that may lead to spurious associations between imputed expression and outcomes. For each gene, we fit two univariate regression models: one adjusting for the top 10 genotype PCs and one adjusting for PCs plus estimated cell fractions. We compared coefficients using the method of Clogg et al.^35^.

## Results

### Overview of the proposed framework

We proposed CausalCellInfer, a framework which integrates scI-GCM with a deep neural network to deconvolute cell proportions and employs a regularized matrix completion algorithm (ENIGMA) to estimate the corresponding CTS profiles. CausalCellInfer can reliably and efficiently deconvolute cell compositions and estimate the corresponding expression profiles from large-scale bulk expression data, including GWAS-predicted expression.

CausalCellInfer mainly employs scI-GCM to identify markers (genes) for each cell type. scI-GCM is based on the principle of invariant causal prediction, designed to identify the “causal” genes for each cell-type. It then uses these identified markers to train the deep neural network-based cell deconvolution model. Subsequently, it employs ENIGMA to infer the corresponding CTS expression profiles based on the estimated cell proportions.

We verified the efficacy of our proposed method by comparing it against existing state-of-the-art cell deconvolution methods, including NNLS, DWLS, MuSiC, Scaden, and DISSECT across various real and pseudo-bulk samples. Finally, we applied our proposed framework to deconvolute four GWAS-predicted tissue-specific expression profiles in the UKBB, i.e., frontal cortex, pancreas, subcutaneous adipose tissue (SAT) and visceral adipose tissue (VAT), using disease status as environment variable. With the deconvoluted cell compositions and corresponding CTS expression profiles, we conducted a wide range of analyses for 29 clinically relevant phenotypes (4 ‘main’ disorders, 8 prognostic traits pertaining to the 4 main disorders, 17 other related disorders) in the UKBB, including cell type association analysis, cell type-specific TWAS and causal gene detection, and (tissue-level) TWAS controlled for cell proportions.

### Performance of scI-GCM for marker selection and cell-type deconvolution under confounding

To evaluate marker selection under environment-induced confounding, we compared scI-GCM against another causal-screening approach (PC-simple) and three representative state-of-the-art methods (DWLS, MuSiC, and Scaden) using simulated scRNA-seq and bulk transcriptomic data. We considered two scenarios with different confounding effect sizes (modest and substantial) to mimic realistic settings in which factors such as disease status impose strong confounding effects on cell-specific expression profiles.

Despite using the smallest marker gene set, scI-GCM outperformed all competitors in terms of concordance correlation coefficient (CCC), root mean square error (RMSE), and mean absolute error (MAE) in simulations with four cell types (Figure 2). Notably, as the magnitude of confounding increased, the performance advantage of scI-GCM became more pronounced. In contrast, DWLS struggled to deconvolute samples simulated from diseased conditions (Figure 2, the median of CCC, MAE and RMSE is comparable to other methods), although it achieved good performance on control samples. This discrepancy likely arises because DWLS constructs its deconvolution signature matrix from control samples, which limits its generalizability to samples affected by confounding factors such as disease.

**Figure 2.**
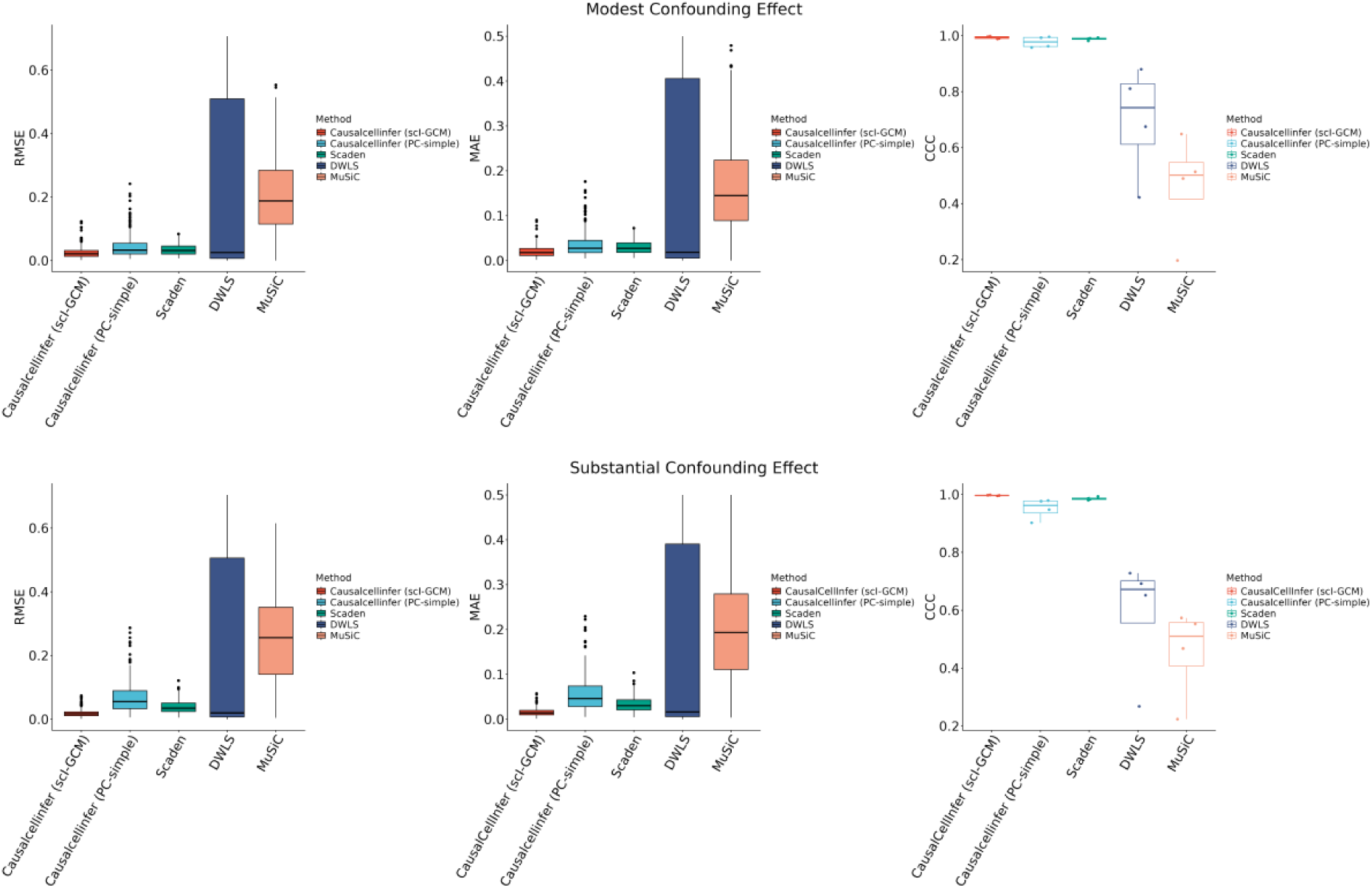
Benchmarking of cell deconvolution accuracy on simulated bulk samples comprising 4 cell types with varied confounding effects.

We also assessed cell marker quality by different algorithms. scI-GCM also demonstrated superior performance in identifying highly ranked differentially expressed genes (DEGs) relative to PC-simple. Genes were ranked by the mean inter-cell-type difference in decreasing order; the top 100 were considered DEGs. Marker gene sets identified by scI-GCM achieved higher PPV for DEGs than those derived from PC-simple (Table 1). Moreover, as confounding effects intensified, scI-GCM more reliably identified trustworthy DEGs for downstream deconvolution.

**Table 1.** Comparison of scI-GCM and PC-simple in identifying DEG genes.

| Scenario | Positive predictive value (PPV) |  |
| --- | --- | --- |
|  | scI-GCM | PC-simple |
| 4 cell types with modest confounding effects | 0.476 | 0.207 |
| 4 cell types with substantial confounding effects | 0.627 | 0.208 |

### Deconvolution accuracy on pseudo-bulk mixtures

We next benchmarked deconvolution accuracy on pseudo-bulk mixtures derived from scRNA-seq references from four tissues (cortex, pancreas, SAT, VAT). The comparative results with existing methodologies are illustrated in Figures 3-4 and Table 2. Our proposed method, CausalCellInfer (utilizing either scI-GCM or PC-simple alone for marker selection), was benchmarked against five state-of-the-art cell deconvolution techniques: NNLS, DWLS, MuSiC, DISSECT and Scaden. NNLS, DWLS, and MuSiC are all signature-based methods that leverage reference scRNA-seq data to generate the gene expression profiles (GEPs) necessary for cell deconvolution. In contrast, Scaden, DISSECT and our method are deep-learning based methods that employ the reference scRNA-seq data for pseudo-bulk training set construction, which is subsequently used to train cell deconvolution models. To mitigate the risk of overfitting, we terminated the training of deep-learning models after 5000 steps, adhering to the Scaden pipeline.

**Figure 3.**
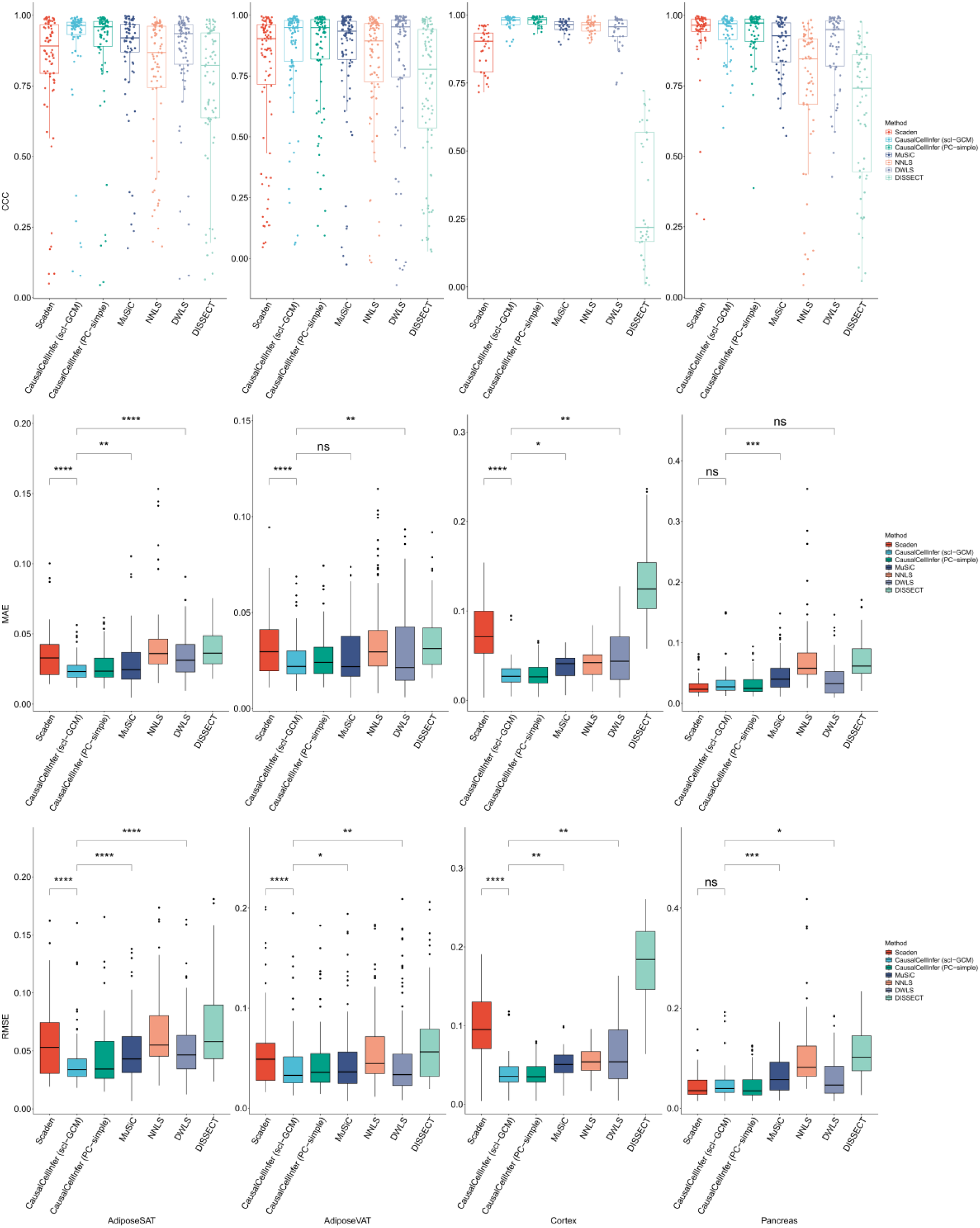
Benchmarking of cell deconvolution accuracy on pseudo-bulk data constructed by combining reference scRNA-seq examples.

**Figure 4.**
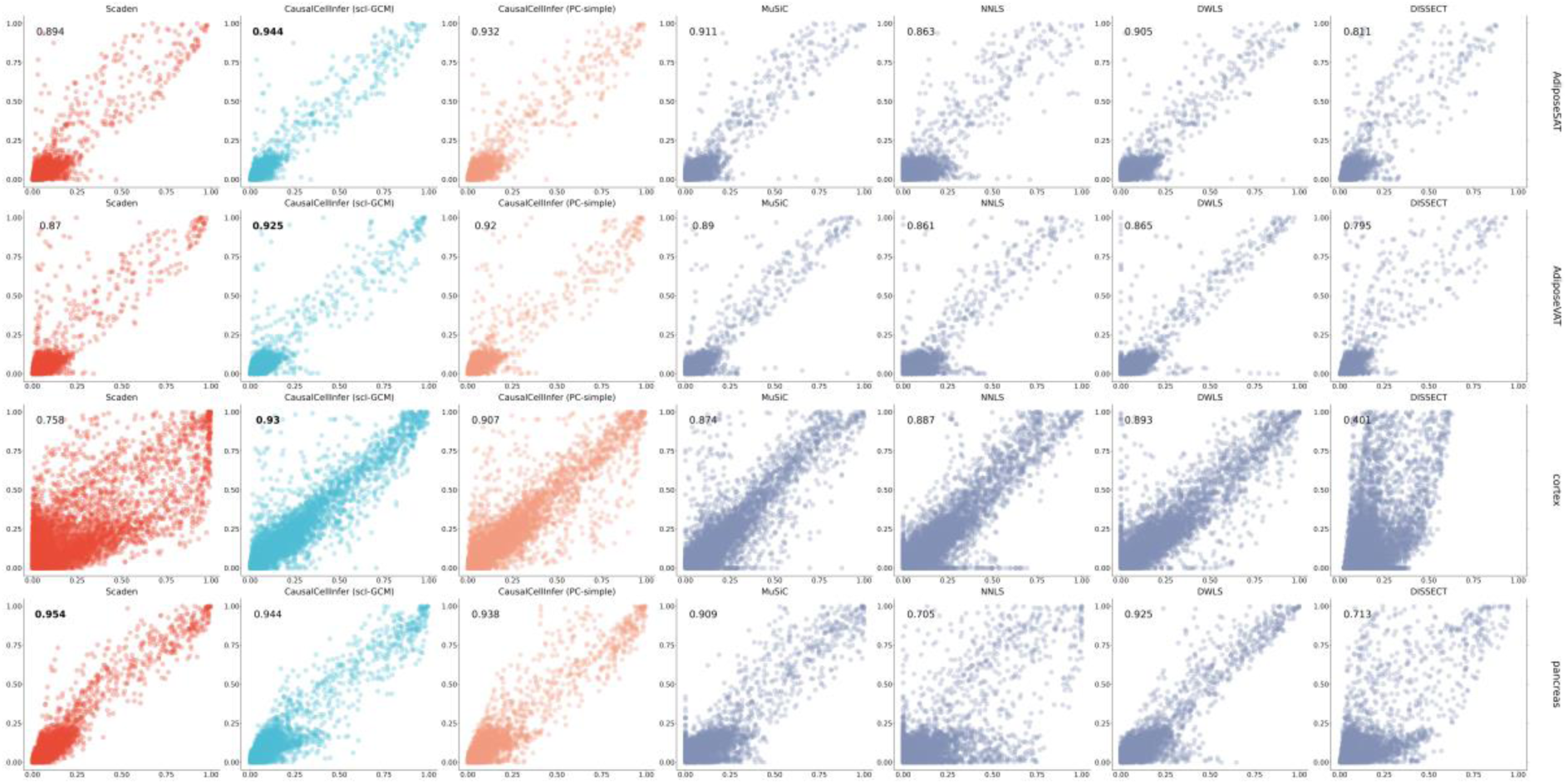
Benchmarking of cell deconvolution accuracy on pseudo-bulk data constructed by combining reference scRNA-seq examples with Lin’s Concordance Correlation Coefficient (CCC).

**Table 2.** Comparison of cell deconvolution accuracy on pseudo-bulk data constructed by combining reference scRNA-seq examples.

| Methods | Cortex |  |  | Adipose SAT |  |  | Adipose VAT |  |  | Pancreas |  |  |
| --- | --- | --- | --- | --- | --- | --- | --- | --- | --- | --- | --- | --- |
|  | CCC | RMSE | MAE | CCC | RMSE | MAE | CCC | RMSE | MAE | CCC | RMSE | MAE |
| Scaden | 0.758 | 0.0964 | 0.0743 | 0.894 | 0.0578 | 0.0347 | 0.870 | 0.0555 | 0.0331 | **<u>0.954</u>** | **<u>0.0448</u>** | **<u>0.0295</u>** |
| CausalCellInfer (scI-GCM) | **0.930** | *0.0422* | *0.0321* | **0.944** | **0.0409** | **0.0248** | **0.925** | **0.0437** | **0.0259** | *0.944* | 0.0508 | 0.0357 |
| CausalCellInfer (PC-simple) | *0.907* | **0.0387** | **0.0295** | *0.932* | *0.0454* | *0.0279* | *0.920* | *0.0459* | *0.0271* | 0.938 | *0.0464* | *0.0314* |
| MuSiC | 0.874 | 0.0508 | 0.0371 | 0.911 | 0.0506 | 0.0299 | 0.890 | 0.0485 | 0.0279 | 0.909 | 0.0664 | 0.0476 |
| NNLS | 0.887 | 0.0535 | 0.0415 | 0.863 | 0.0665 | 0.0429 | 0.861 | 0.0587 | 0.0376 | 0.705 | 0.1104 | 0.0781 |
| DWLS | 0.893 | 0.0633 | 0.0499 | 0.905 | 0.0541 | 0.0337 | 0.865 | 0.0520 | 0.0306 | 0.925 | 0.0633 | 0.0423 |
| DISSECT | 0.401 | 0.1775 | 0.1287 | 0.811 | 0.0705 | 0.0389 | 0.795 | 0.0656 | 0.0357 | 0.713 | 0.1128 | 0.0736 |

The top 2 performing methods are underlined. The best performing method is also bolded, while the 2<sup>nd</sup> best methods are in italics.
Note: CCC is the abbreviation for Lin's Concordance Correlation Coefficient, RMSE is the abbreviation for Root Mean Square Error, MAE is the abbreviation for Mean Absolute Error.

In summary, our proposed method, CausalCellInfer demonstrated consistent superiority over existing methods such as NNLS, MuSiC, DWLS, DISSECT and Scaden, in deconvoluting pseudo-bulk RNA-seq data. Notably, our approach achieved satisfactory Concordance Correlation Coefficient (CCC), Root Mean Square Error (RMSE) and Mean Absolute Error (MAE) across the four pseudo-bulk scenarios, when compared to other cell deconvolution methods.

Considering RMSE and MAE, CausalCellInfer(scI-GCM) was the best performing method in two out of the four tissues tested (SAT, VAT); CausalCellInfer (PC-simple) performed the best for cortex, yet CausalCellInfer(scI-GCM) ranked second with very close performance. For pancreas, Scaden performed the best, however CausalCellInfer (both scI-GCM and PC-simple) ranked 2^nd^ and 3^rd^ with close performance. Considering the performance according to CCC, CausalCellInfer(scI-GCM) was the best for three tissues (cortex/SAT/VAT). For pancreas, Scaden again led based on CCC, followed closely by CausalCellInfer(scI-GCM) and CausalCellInfer(PC-simple). Taken together, our proposed framework achieves top performance in cell type deconvolution.

Both Scaden and our proposed method employ DNNs for cell deconvolution. However, our method consistently outperforms Scaden, as evidenced by significantly CCCs and lower MAE and RMSE across most test scenarios. Moreover, our method exhibits a markedly smaller variance in CCCs across all cell types (Figure 3), underscoring its robustness in deconvoluting cell compositions. During the training phase of the deconvolution model, we opted to use selected marker genes identified by scI-GCM/PC-simple, as opposed to all highly variable genes. Our results indicate that focusing on a subset of informative marker genes that are stable across environments can significantly enhance performance within a deep learning framework. This approach likely leads to a less complex, more robust and transferable deconvolution model less affected by noise in the data.

#### Evaluation on real-world datasets

In the evaluation of real-world datasets, our method exhibited comparable performance to Scaden, as indicated by analogous RMSE and MAE values. Importantly, our methodology outperformed alternative approaches by attaining a superior CCC across most cell types. Upon application to the Monaco and GSE107572 dataset, our proposed method (CausalCellInfer(scI-GCM)) excelled, surpassing all other methods in terms of both MAE and RMSE. Furthermore, it achieved the highest average CCC among all evaluated methods (Figure 5). Interestingly, This pattern is consistent with prior benchmarking on the same dataset^17^, where MuSiC underperformed in real-world PBMC dataset.

**Figure 5.**
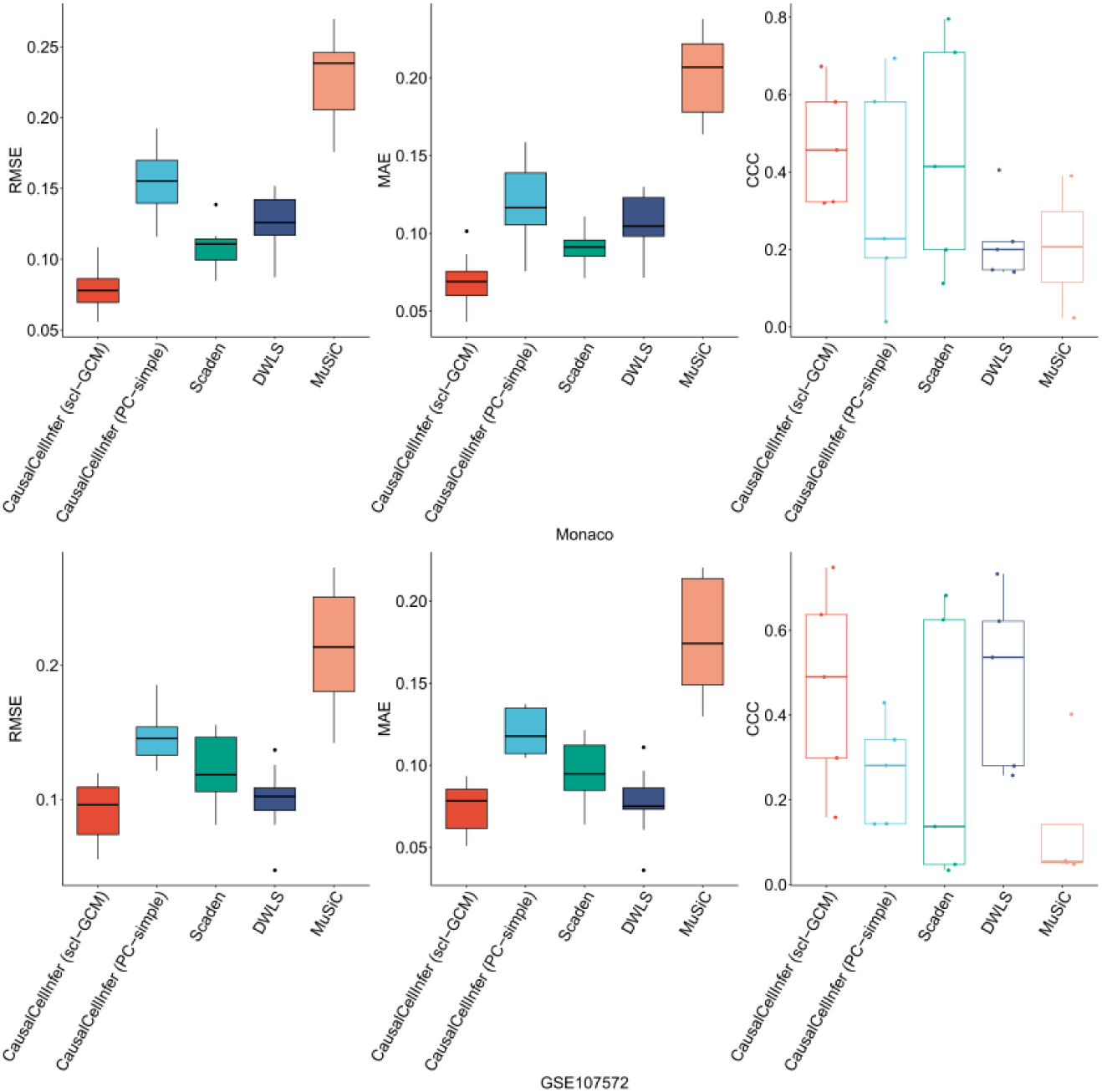
Benchmarking of cell deconvolution accuracy on real bulk data

### Validity of inferred cell-type specific expression profile

We conducted a comparison analysis on the cell-type specific DEGs identified from our estimated CTS profiles (from GWAS data) and the reference scRNA-seq datasets for similar phenotypes. If the inferred CTS expression profiles are reliable, we expect a significant overlap between the DEG identified from the two types of datasets. Here we regard the DEGs from the actual scRNA-seq data as the reference for comparison.

We reveal that the DEGs identified via the CTS expression profiles based on our method and GWAS data exhibit substantial overlap with satisfactory PPV for the majority of cell types. For instance, the DEGs identified for primary cell types in the pancreas, namely acinar cells, alpha cells, beta cells, and delta cells, achieved a PPV exceeding or around 0.5, significantly higher than expectation (Table S1). In adipose tissue, the two predominant cell types, adipocytes and adipose-derived stem and progenitor Cells (ASPCs), demonstrated the highest PPV across all cell types, with values greater than 0.8 in SAT and 0.6 in VAT. These findings validate the reliability of our proposed method for inferring CTS expression profiles.

### UKBB application: cell-type proportion and cell-type specific expression estimation at biobank scale

We applied CausalCellInfer to PrediXcan-imputed tissue-specific expression for ∼500,000 UKBB participants, estimating cell-type proportions and CTS expression profiles for depression (frontal cortex), type 2 diabetes (pancreas), and obesity (SAT and VAT). Binary traits (except obesity) were defined using self-reported diagnoses and ICD codes. Obesity was defined as BMI > 30.

### Distinguishing cell types associated with various diseases

To elucidate the relationship between the estimated cell proportions and diseases/traits, we conducted univariate association tests for each cell type via logistic/linear regression, with the trait/disease of interest as outcome. We revealed that the estimated proportions of various cell types were associated with different diseases, as detailed below.

#### Depression

We deconvoluted the frontal cortex into six distinct cell types and discovered that the estimated cell proportions for astrocytes, micro/macro cells, and oligodendrocytes were significantly associated with risk of depression (Figure 6, Table S2). Notably, a lower inferred proportion of microglial/macrophage cells and oligodendrocytes were associated with depression, while the proportion of neurons was elevated. These observations are consistent with existing literature; for instance, Rajkowska et al.^36^ observed a decrease in both the density and ultrastructure of oligodendrocytes in the prefrontal cortex in depression.

**Figure 6.**
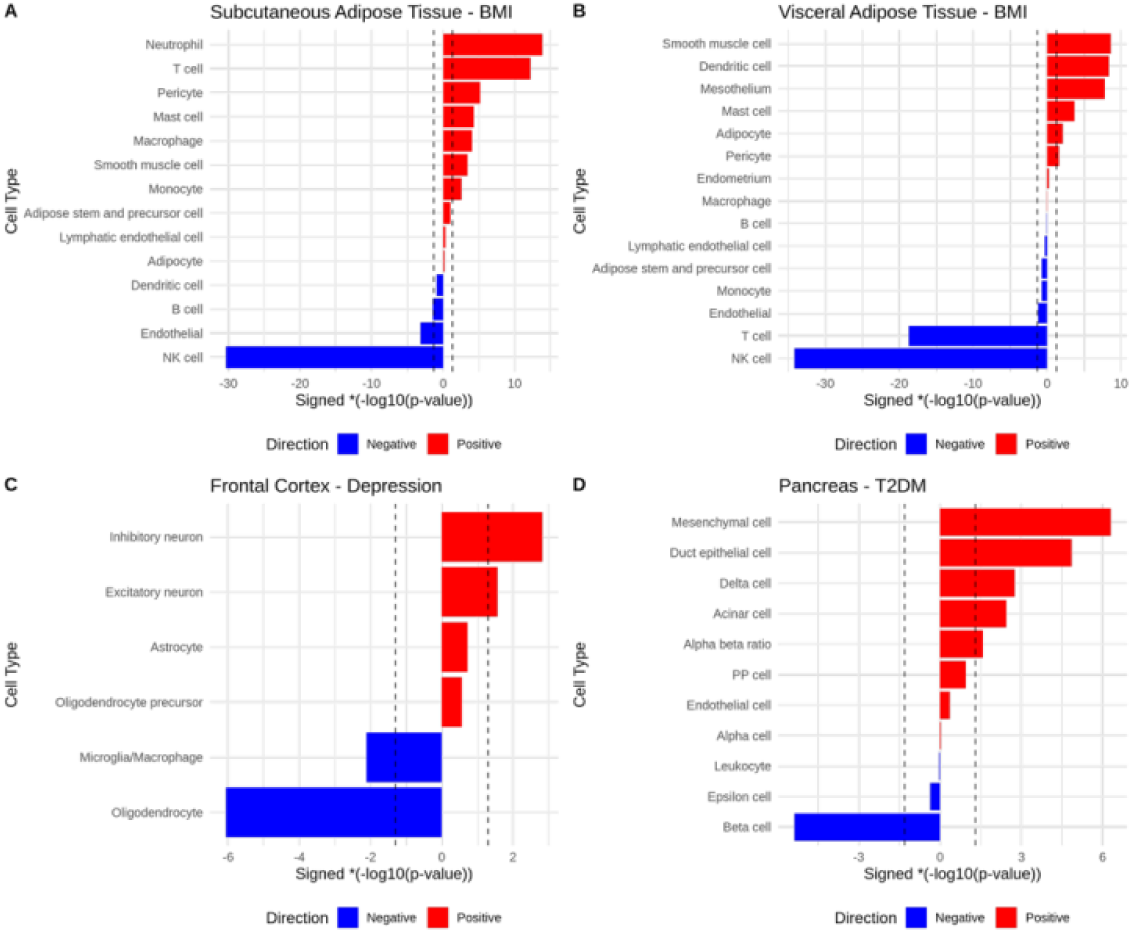
Associations between estimated cell proportions and disease onset

#### Type 2 Diabetes

In our analysis of ten distinct cell types within the pancreas, we observed that the inferred proportions of six cell types, including beta cells, exhibited a significant association with T2DM risks. Notably, significantly lower inferred proportions of beta cells were observed in T2DM. Conversely, we detected associations of elevated proportions of acinar, delta, duct epithelial, and mesenchymal cells with T2DM, with the increase in mesenchymal cell proportions being especially prominent (Figure 6). We also observed significant alpha beta ratio increase in T2DM patients. Numerous studies have underscored the link between T2DM and a decrease in beta-cell mass and function^37,38^. Also, previous GWAS have pinpointed *IRX3* as a susceptibility gene for T2DM, and knockdown of the *IRX3* orthologue in zebrafish led to a decrease in insulin-producing beta cells^39^.

#### Obesity

In subcutaneous and visceral adipose tissues (SAT/VAT), we deconvoluted 14 and 15 distinct cell types respectively, and tested associations with obesity status. More than half of these cell types, including smooth muscle cells (SMC), and adipocyte, were found to be associated with obesity. Interestingly, the involvement of immune-related cell types in obesity was observed to vary between adipose tissues. Some cell types, such as natural killer cells (NK cells) and T cells, were associated with obesity in both tissues (SAT and VAT), while others, like neutrophils and macrophage, were associated with obesity in only one tissue.

Compared to normal subjects, there was a significant increase in the estimated proportions of various cell types, including SMCs, macrophage, mast, monocyte, neutrophil, pericyte and T cells in SAT. Conversely, a significant decrease in NK, endothelial and B cell proportion was noted.

Notably, a significant increase in pericyte proportions was also observed in both adipose tissues. Supporting our findings, Tang et al.^40^ demonstrated that pericytes are one of the precursor cells of adipocytes, underscoring their crucial role in obesity development. Watanabe et al.^41^ also showed that mice fed with a high-fat diet (HFD) exhibit a higher level of pericyte count than those on a chow diet.

#### Interactions between cell types and their associations with disease

We tested the effects of cell type interactions on 4 tissue-specific outcome variables using multivariate regression analyses, including depression in the frontal cortex, T2DM in the pancreas, BMI in the SAT, and BMI in the VAT.

Our results revealed that interactions between certain cell types were significantly associated with disease risks (Table 3 and S3). For example, the interaction between astrocytes and microglia/macrophages was linked to depression (based on expression from the frontal cortex). This is consistent with the literature suggesting a disrupted structural communication between excitatory neuron and oligodendrocyte in the frontal cortex, which contributes to both cognitive retardation and emotional instability commonly seen in depression ^42^.

**Table 3.**
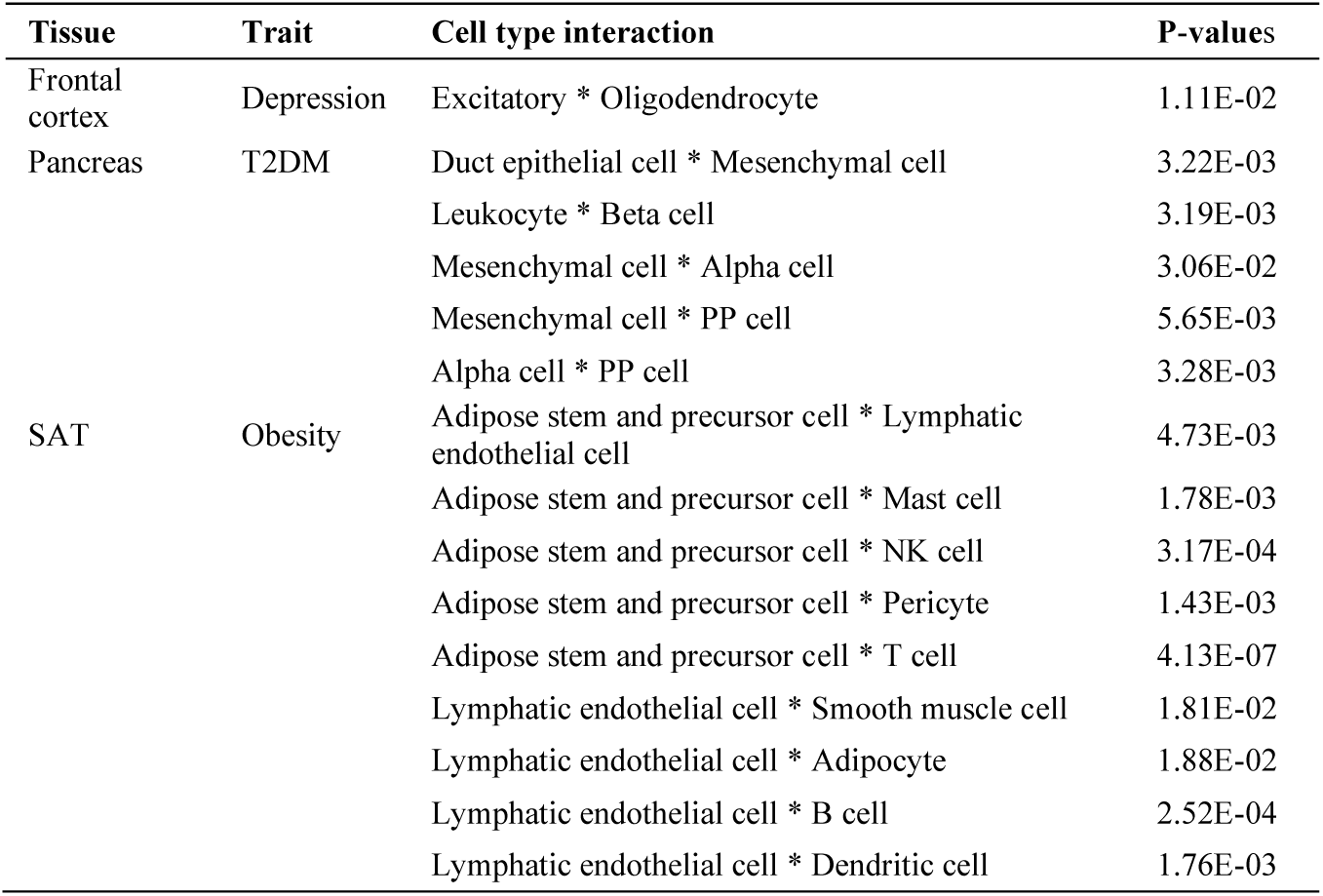
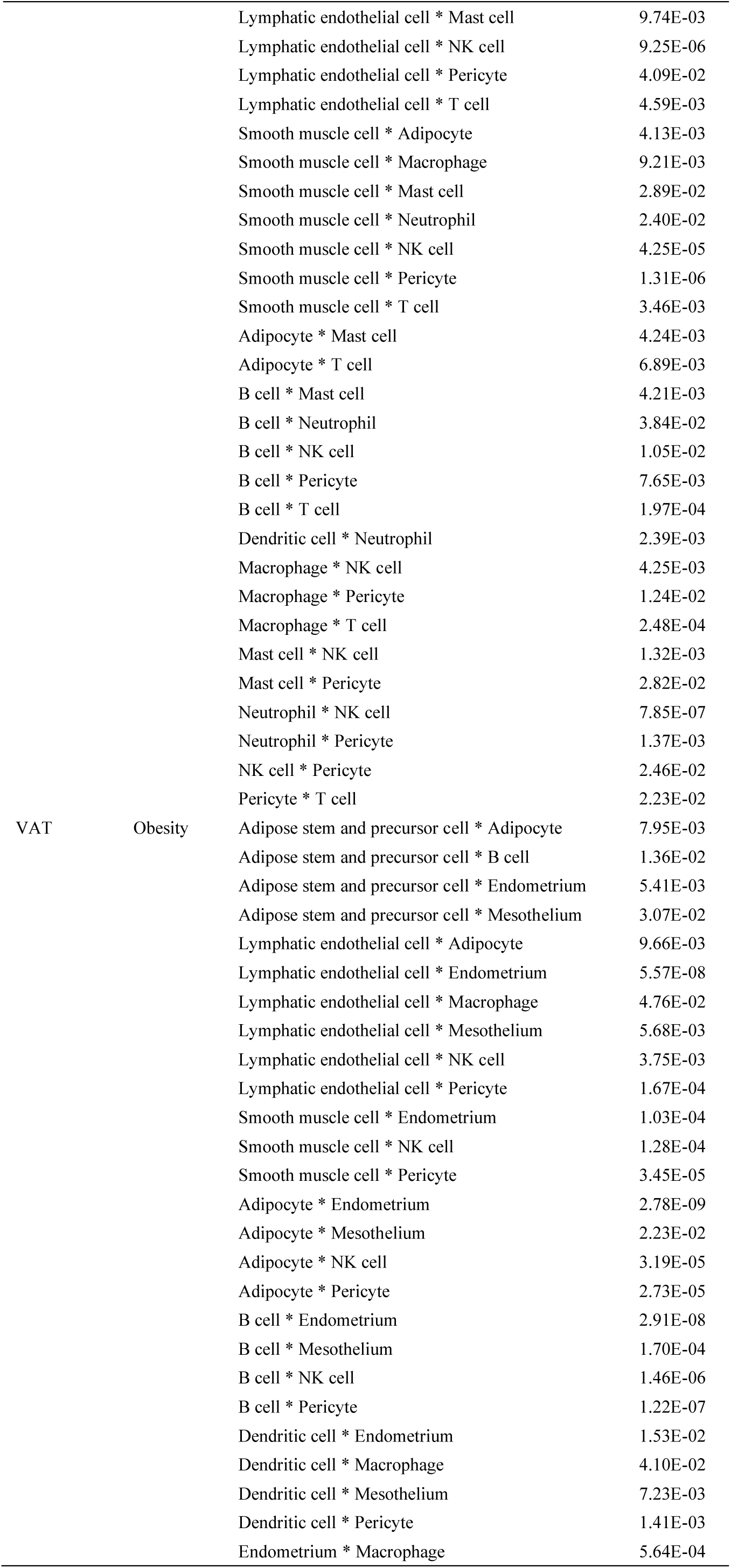
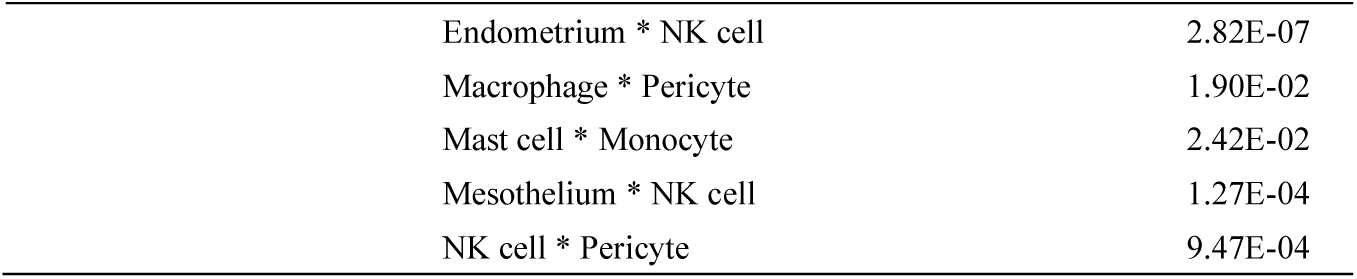
Cell type interactions significantly associated with diseases.

In addition, we found that interaction of cell types *across different tissues* may contribute to disease risks. For example, the interaction between B cells on SAT and dendritic cells in VAT was significantly associated with obesity. Both cell types play crucial roles in maintaining immune homeostasis, and previous studies^43,44^ has revealed significant involvement of inflammation in obesity. To conclude, interactions between certain cell types may play an important role in disease pathogenesis.

### Cell type-specific genes linked to disease susceptibility (focusing on putative direct casual genes as prioritized by PC-simple)

Putative direct causal effect genes for disease susceptibility were found to exhibit variation across different cell types (Figure 7, Table S4). We refer to genes selected by PC-simple as putative direct causal genes. We define genes detected in exactly one cell type as cell-type-specific and genes detected in ≥2 cell types as cell-type-common.

**Figure 7.**
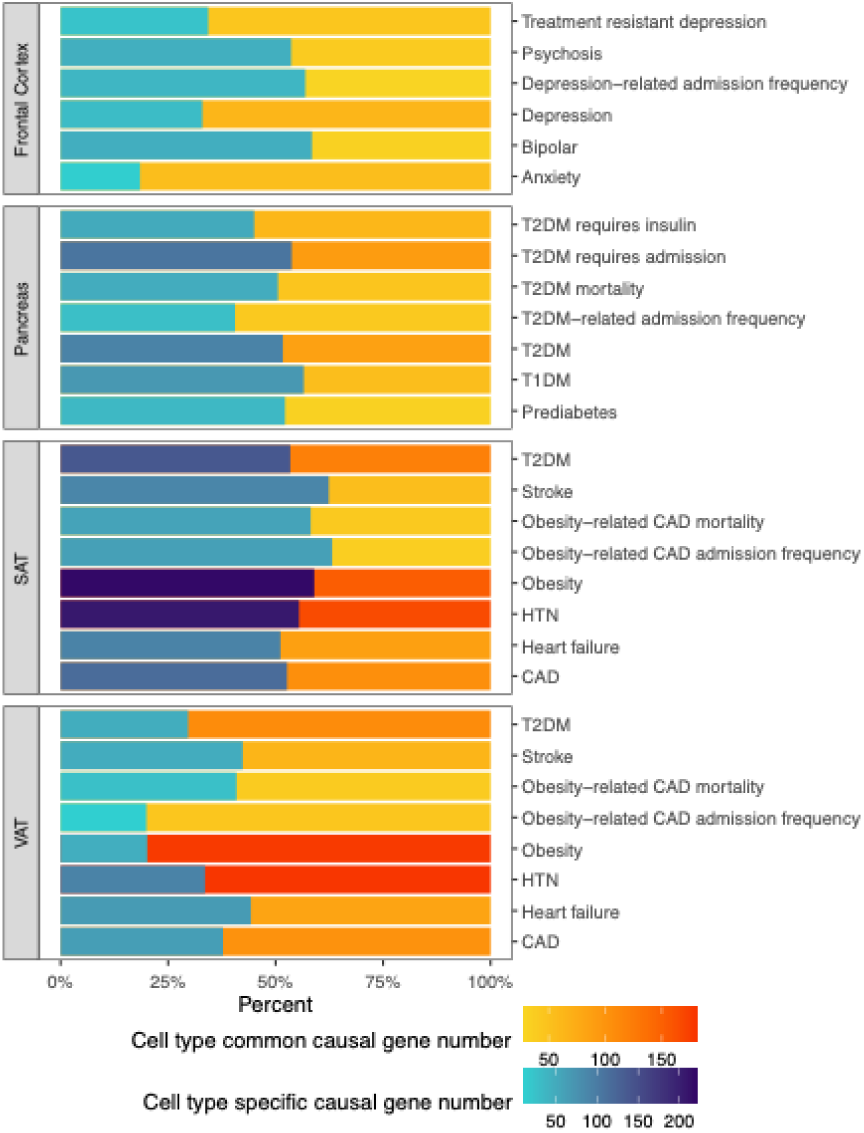
Cell specificity comparison of the identified putative direct causal genes by PC-simple for disease onset and prognoses

#### Depression

For depression, approximately one-third of the 94 prioritized genes were cell-type-specific. A notable example is *KLC3*, identified as a causal gene for depression susceptibility exclusively within excitatory neurons (Table 4; Table S4). This finding aligns with a previous study^45^ reporting neuronal hypomethylated changes of *KLC3* in the prefrontal cortex of bipolar patients, suggesting a potential epigenetic link to mood disorders.

**Table 4.**
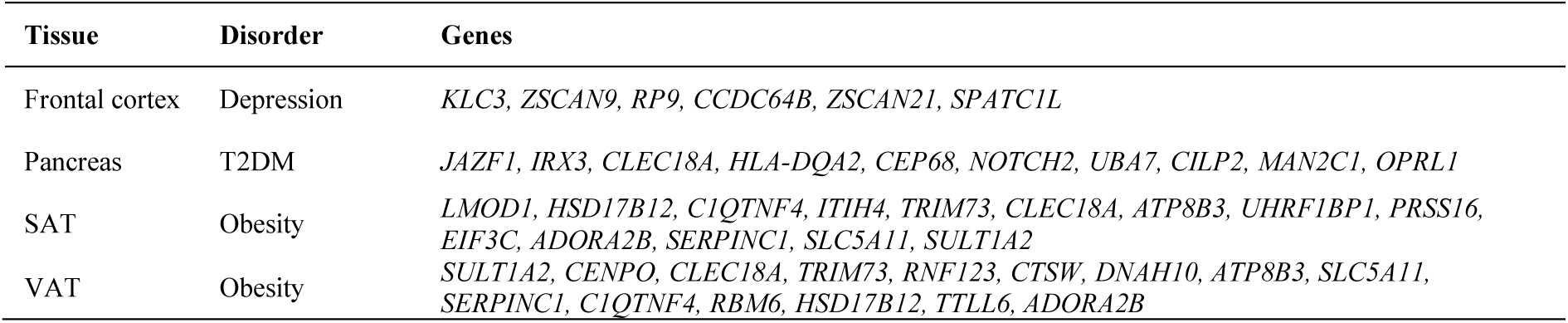
Top putative direct causal genes by PC-simple (ranked by P-value) from inferred cell-type specific (CTS) expression profiles.

Our analysis revealed a nuanced relationship between cell-type alterations and disease status. Some cell types exhibited both altered proportions and changes in gene expression, while others showed only gene expression alterations without significant proportional changes. For instance, despite no significant association between oligodendrocytes precursor cells (OPCs) proportion and depression susceptibility, this cell type harbored 16 putative direct-effect genes.

#### T2DM

For T2DM, we identified 178 putative direct-effect genes associated with disease risk, with varying involvement across different cell types. In contrast to depression, where cell-type specificity was more prominent, nearly half (86 genes, ∼48.3%) of the T2DM-associated genes were common, detected in two or more cell types (Figure 7, Table S4). For instance, *JAZF1* was identified as a direct causal gene in 6 distinct cell types, such as acinar, alpha, beta, and delta cells (Table S4). The association of *JAZF1* with increased T2DM risk is well-established^46,47^.

#### Obesity

Our analysis of obesity revealed a substantial number of putative direct-effect genes in both subcutaneous and visceral adipose tissues, but with a large difference in the distribution of cell-type-specific versus common genes. In SAT, a high degree of cell-type specificity was observed, with approximately 69.7% of causal genes being specific to a single cell type (only 77 genes, or 30.3% of ∼254 identified genes, were cell-type-common). Conversely, VAT displayed a much higher proportion of common genes, with 178 genes (79.8% of ∼223 identified genes) contributing to obesity across multiple cell types. This suggests a broader, multi-cellular involvement of causal mechanisms in VAT.

A notable cell-type-specific example in SAT is *PRSS53* (also known as *POL3S*), identified as a causal gene exclusively in adipocytes. This finding is supported by a previous study^48^ showing *PRSS53* downregulation in obesity. Given *PRSS53*’s critical role in maintaining mitochondrial health and function for metabolism^49^, its potential causal role in obesity development within SAT is further substantiated.

### Cell type-specific genes involved in disease prognosis (focusing on putative direct casual genes as prioritized by PC-simple)

Identifying specific genes that contribute to disease outcomes or prognosis is crucial for developing personalized treatments. Leveraging detailed prognosis data from the UKBB, which are often absent in single-cell studies, we conducted analyses to pinpoint putative direct-effect genes associated with disease-specific mortality and hospitalization frequencies.

#### Depression

For depression, our analysis revealed distinct patterns for treatment resistance and hospitalization frequency. A substantial proportion (∼65.7%) of putative direct-effect genes linked to treatment resistance were common, identified in at least two cell types (Table S4.a). Conversely, genes associated with the frequency of hospital admissions were predominantly cell-type specific (∼56.9%) (Table S4.a).

#### Type 2 Diabetes Mellitus

In T2DM, we observed that a majority (∼59.4%) of putative direct-effect genes influencing hospitalization frequencies were common across multiple cell types (Table S4.b). However, for T2DM-related mortality, a slightly different distribution emerged, with approximately half (∼49.5%) of the causal genes being cell-type common (Table S4.b). For example, we identified the gene *LIMK1* as a determinant of mortality in T2DM within beta cells. This finding aligns with a recent research indicating a relationship between genetic polymorphisms of *LIMK1* and glucose dysregulation and insulin exocytosis^50^.

#### Obesity

Given obesity’s established role as a risk factor for coronary artery disease (CAD)^51^, we investigated genes influencing CAD-related mortality and hospitalization frequency in obese patients. A notable divergence was observed between adipose tissues: SAT showed a higher proportion of cell-type-specific genes, while VAT exhibited a higher proportion of common genes influencing these outcomes (Table S4.c and Table S4.d).

#### Other Disorders

The distribution of identified causal genes varied considerably across tissues. In SAT, for most obesity-related traits (CAD, T2DM, hypertension, heart failure, stroke), cell-type-specific causal genes predominated. The inverse pattern was consistently found for these same traits in VAT.

Similarly, for traits associated with the frontal cortex (anxiety, bipolar disorder, psychosis) and pancreas (prediabetes, T1DM, T2DM requires admission, T2DM requires insulin), the majority of identified causal genes were found to be cell-type specific.

### OpenTargets enrichment results

We conducted enrichment analyses for depression, T2DM and obesity. For obesity, we performed four sets of enrichment analyses for each pair of tissues associated with obesity. Specifically, we investigated whether the genes we identified were significantly overrepresented as targets for BMI/obesity/weight/WHR (waist-hip-ratio) in the OpenTargets database.

Encouragingly, the sets of causal genes identified for all the studied diseases were significantly enriched among the disease targets catalogued in OpenTargets (Table 5). This observation provides further validation for the usefulness of our proposed framework in estimating cell proportions and CTS expression profiles.

**Table 5.**
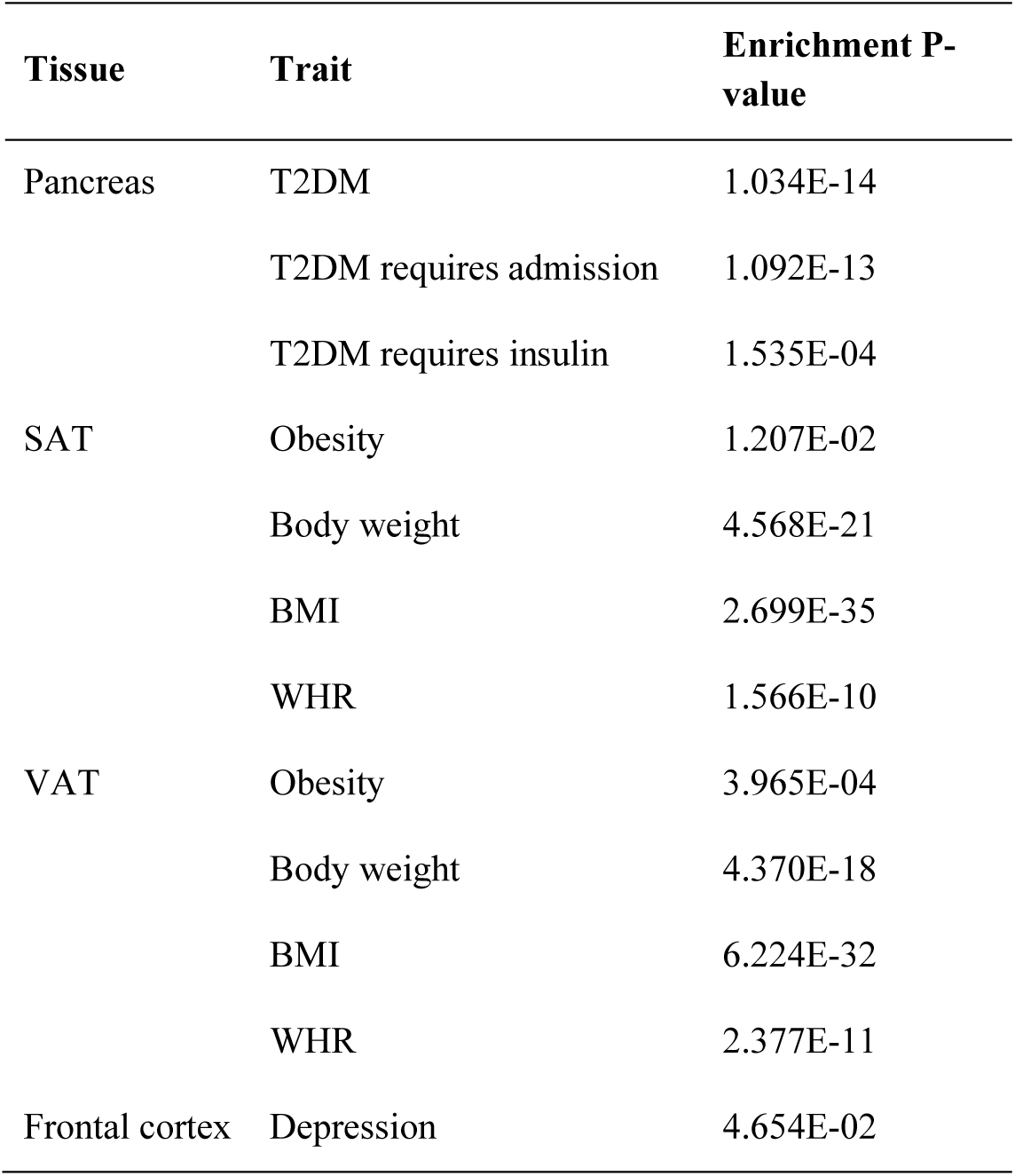
OpenTargets enrichment results based on gene-sets identified for disease susceptibility.

| Tissue | Trait | Enrichment P-value |
| --- | --- | --- |
| Pancreas | T2DM | 1.034E-14 |
|  | T2DM requires admission | 1.092E-13 |
|  | T2DM requires insulin | 1.535E-04 |
| SAT | Obesity | 1.207E-02 |
|  | Body weight | 4.568E-21 |
|  | BMI | 2.699E-35 |
|  | WHR | 1.566E-10 |
| VAT | Obesity | 3.965E-04 |
|  | Body weight | 4.370E-18 |
|  | BMI | 6.224E-32 |
|  | WHR | 2.377E-11 |
| Frontal cortex | Depression | 4.654E-02 |

### Results of further pathway enrichment analyses

We combined the identified cell-type specific causal genes ((prioritized by the PC-simple algorithm) for pathway enrichment analysis. The enriched pathways were shown in Table S5. For instance, “Pyrophosphate hydrolysis” was significantly enriched for depression, consistent with Sha et al.^52^ work suggesting its protective role in astrocytes against depression via lysosomal function.

For T2DM, pathways like “Vitamin B3 (nicotinate and nicotinamide) metabolism”, “Pre-NOTCH Processing in the Endoplasmic Reticulum”, and “Pre-NOTCH Processing in Golgi T2DM” were significantly enriched. This aligns with Lu et al.^53^ findings on dysregulated Notch signaling worsening T2DM through hepatic fat deposition and glucogenesis.

We also identified enriched pathways based on gene sets associated with disease prognosis (Table S5). For instance, the “Apoptotic cleavage of cellular proteins” pathway was significantly enriched in treatment-resistant depression (TRD). This aligns with Kosten et al.^54^ who showed that antidepressant treatment can differentially regulate apoptotic genes in the brain structures implicated in the etiology and treatment of depressive disorder. Fries et al.^55^ also showed that apoptotic events may be responsible for the chronic low-grade inflammation in depressive patients. The “Ceramide signaling pathway” pathway was enriched in relation to the frequency of depression-related admissions. Gulbins et al.^56^ reported that increased ceramide concentration can lead to the reversal of stress-induced MDD via activation of autophagy.

“Role of mitochondria in apoptotic signaling” emerged as a significantly enriched pathway in relation to T2DM-related mortality, supporting existing literature that apoptotic signaling drives cell death in diabetic complications like cardiovascular diseases (CVD) ^57^ and beta cell failure^58^, increasing T2DM mortality.

### Results from conditional TWAS (controlled for cell-type proportions)

We compared trait-associated genes identified from two different linear models using bulk expression data, one with estimated cell proportions as covariates (conditional TWAS) and one without. Genes from the adjusted TWAS are shown in Table 6 and S6. Adjusting for estimated cell proportions when performing TWAS analysis led to significant coefficient changes for numerous genes (Table 7 and S7-8), consistent with cell composition acting as a confounder or mediator of bulk-level associations. For example, adjusting for cell proportions resulted in significant effect size changes for ∼59.7% of T1DM-associated genes. Some genes were significant only in the standard TWAS (e.g., *EFEMP2* in SAT obesity), illustrating that bulk-level associations can be sensitive to compositional variation.

**Table 6.** Examples of top (ranked by P value) trait-associated genes identified from TWAS after adjustment for estimated cell-type proportions.

| Tissue | Disorder | Genes |
| --- | --- | --- |
| Frontal cortex | Depression | *ZSCAN9, BTN3A2, C4A* |
| Pancreas | T2DM | *IRX3, NOTCH4, CD84* |
| SAT | Obesity | *RTN4, TPCN2, SEMA3F* |
| VAT | Obesity | *TBL2, NOVA1, SULT1A2* |

**Table 7.**
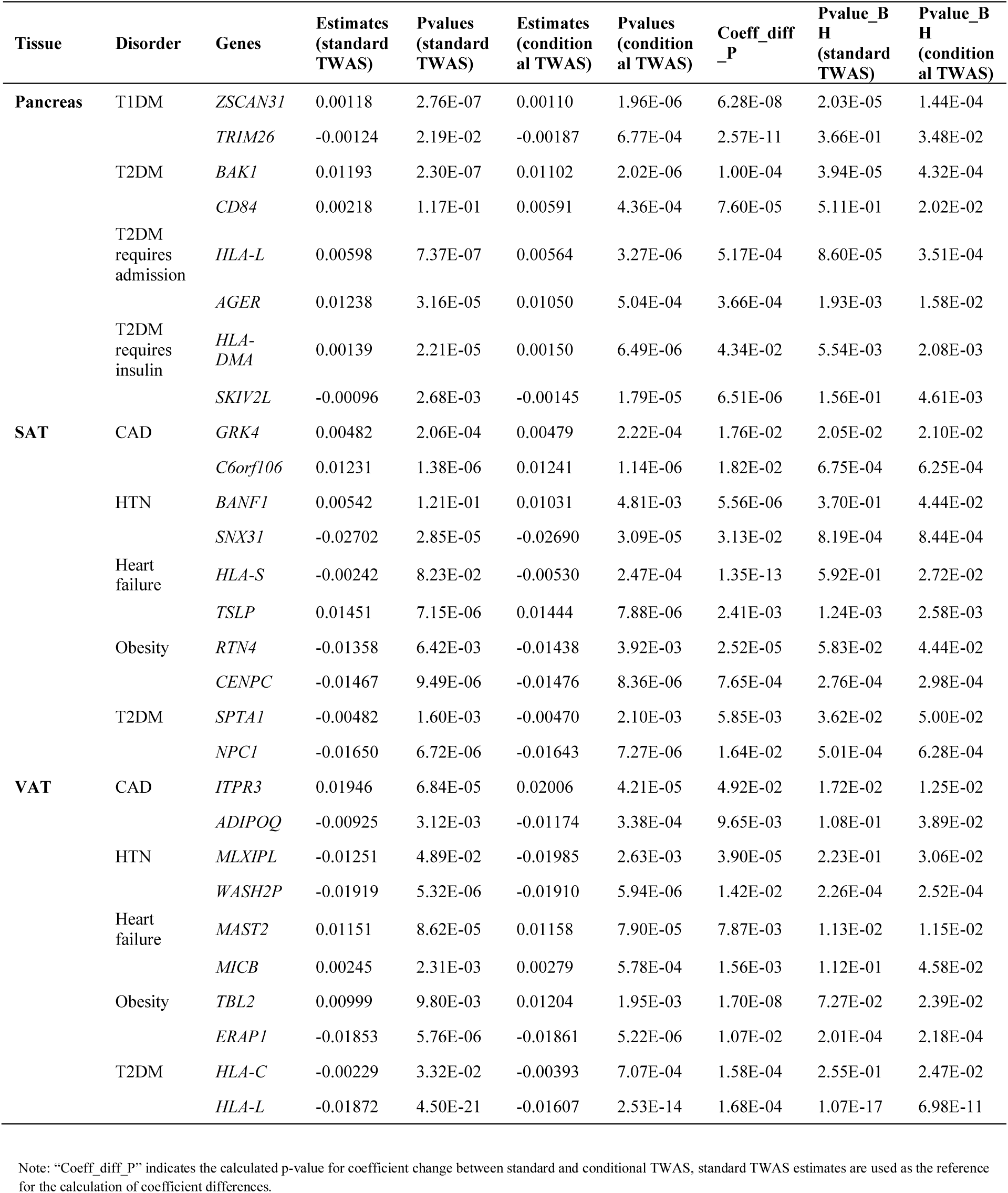
Trait-associated genes from a standard TWAS analysis with significant coefficient changes after adjusting for estimated cell-type proportions.

### Results from CTS-TWAS (univariate association tests)

Here we report the results from CTS-TWAS based on univariate association tests (distinct from PC-simple, which involves repeated conditional tests). Genes identified in at least two cell types were classified as ‘cell-type common’, while those in only one were ‘cell-type specific’.

Similar to direct causal gene analysis by PC-simple, the majority of genes identified from CTS-TWAS were cell-type specific (Table 8, S9-10). The number of significant genes for each trait are listed in Table S9, with FDR<0.05 considered as ‘significant’ genes. Among traits with significant genes, T2DM-related traits typically showed higher cell type-specificity, whereas most obesity-related traits showed broader sharing across cell types.

**Table 8.**
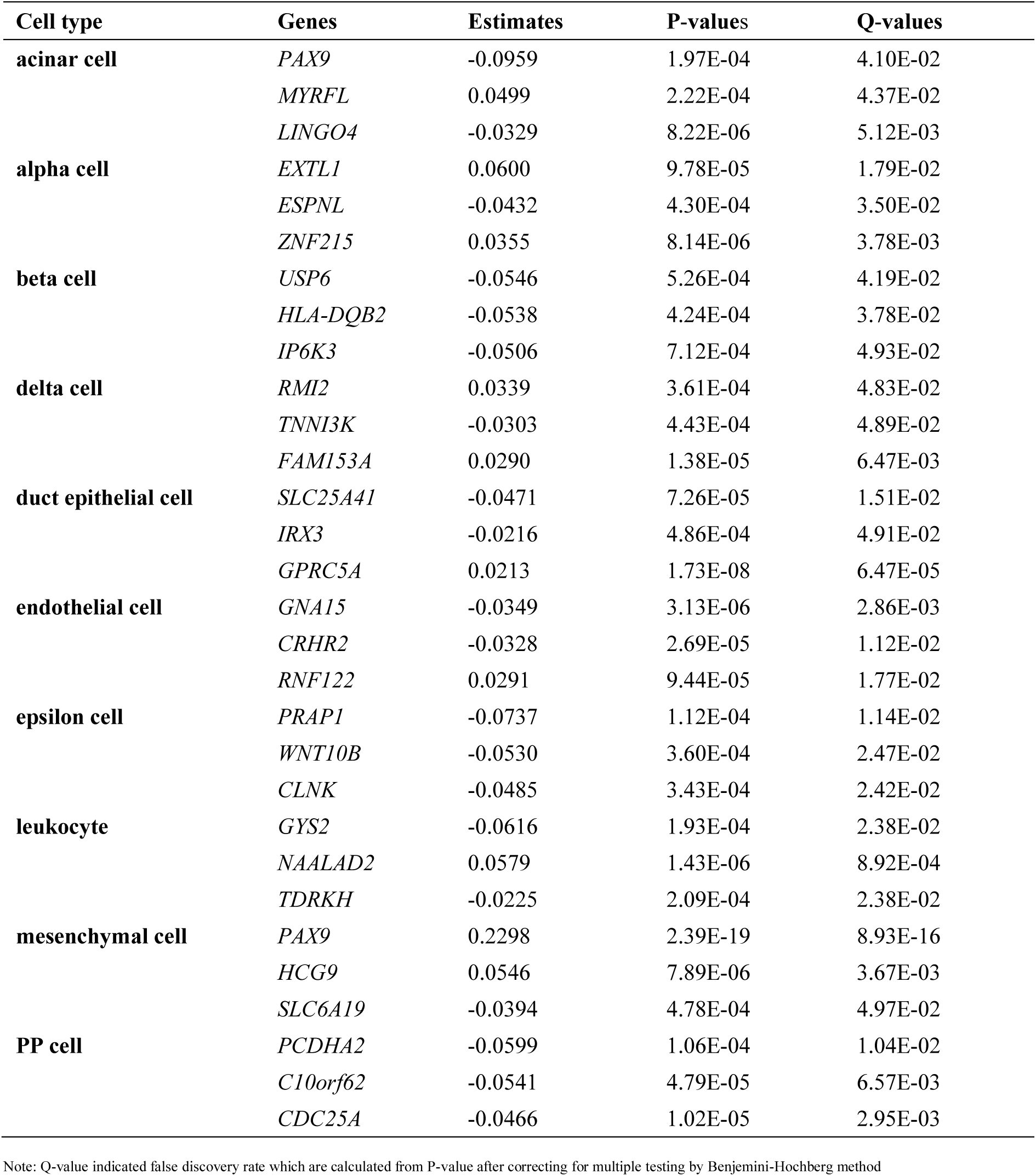
Top-ranked (by absolute effect size from linear regression) trait-associated genes for T2DM (tissue studied: pancreas) identified from CTS-TWAS(univariate tests)

OpenTargets enrichment analysis of CTS-TWAS genes for 4 tissue-disorder pair with >=5 genes (including T2DM and hospitalization frequency (pancreas), obesity (SAT/VAT)) confirmed significant or nominal enrichment among known disease targets listed in OpenTargets (Table 9).

**Table 9.** OpenTargets enrichment results based on gene-sets identified from CTS-TWAS using inferred CTS expression profiles.

| Tissue | Trait | P-value |
| --- | --- | --- |
| Pancreas | T2DM | 9.405E-19 |
|  | T2DM requires admission | 6.044E-19 |
|  | T2DM requires insulin | 7.659E-03 |
| SAT | Obesity | 1.514E-64 |
|  | Body weight | 9.696E-42 |
|  | BMI | 3.201E-60 |
|  | WHR | 9.026E-28 |
| VAT | Obesity | 1.639E-98 |
|  | Body weight | 6.682E-54 |
|  | BMI | 4.919E-63 |
|  | WHR | 1.559E-38 |
| Cortex | Depression | 1.204E-02 |

Furthermore, we compared the genes identified from a causal inference approach (PC-simple, which prioritizes putative direct-effect genes) and univariate association analysis (TWAS) using inferred CTS expression profiles. For most traits, we identified considerably more associated genes from univariate TWAS (Table S9). As PC-simple relies on repeated conditional independence testing, it is more stringent than univariate testing. Interestingly, PC-simple also uncovered novel associations not found in standard association tests (Table S11). We hypothesize that some TWAS-identified genes may be “peripheral” (indirect causal) genes acting through the direct causal genes identified by PC-simple. As expected, if we relax the multiple testing correction criteria, there is an increased overlap between PC-simple and standard association analyses (Table S11).

## Discussion

In this study, we introduced CausalCellInfer, a comprehensive framework that integrates invariant causal prediction, deep learning, and regularized matrix completion, with applications to GWAS-imputed transcriptome profiles. This framework is designed to deconvolute cell compositions and estimate CTS expression profiles from bulk and genotype-imputed transcriptomes at biobank scale (*N*∼500K). As a fully data-driven method, CausalCellInfer first identifies marker genes from the reference scRNA-seq dataset, leveraging the invariant causal prediction principle. It then trains a deep neural network-based deconvolution model using these identified marker genes. Finally, it employs a regularized matrix completion algorithm, ENIGMA, to estimate the corresponding CTS profiles.

We validated the robustness and accuracy of CausalCellInfer in cell deconvolution by comparing it against five state-of-the-art deconvolution methods, namely DWLS, NNLS, Scaden, DISSECT and MuSiC. This comparison was conducted on four simulated and two real peripheral blood mononuclear cell (PBMC) datasets. To further demonstrate the reliability of CausalCellInfer in estimating CTS profiles, we have conducted a wide range of enrichment analyses, including DEGs enrichment analysis, OpenTargets enrichment analysis, and pathway analyses.

Moreover, we applied CausalCellInfer to GWAS-imputed expression datasets from the UKBB (four different tissues) and performed cell-type-level analyses for 29 clinical phenotypes, including susceptibility, prognosis-related outcomes, interaction analyses between cell types, CTS-TWAS, and composition-adjusted (conditional) TWAS. These applications illustrate a central advantage of the approach: it enables cell-type-resolved hypothesis testing in cohorts where the scale and phenotypic depth are far beyond what is typically feasible for single-cell studies.

### Strengths of the framework

The framework presented in this study exhibits several strengths. Firstly, a notable advantage is that CausalCellInfer incorporated a novel marker-gene informed and deep learning-based cell deconvolution model. Marker genes are learned from scRNA-seq reference data rather than relying on predefined marker lists.

Secondly, our framework does not require a constant marker-gene expression assumption as required in classical signature-based methods (i.e., a given marker gene is expressed at a constant level across samples within a specific cell type) for accurate estimation, making it more robust to inter-subject heterogeneities or other technical variabilities across samples.

Thirdly, our method is likely more resistant to the influence of confounders and selection bias than signature-based methods. As also reported in ref^16^, deep learning methods are able to learn latent representations of cell-types. As such, the network nodes effectively capture higher-order features that are less susceptible to noise and technical bias present in the original input expression data.

Fourth, our incorporation of the invariance principle further enhances the robustness of the method to confounding and improves generalizability to new samples.

Fifth, despite being based on deep neural networks, our method is time- and resource-efficient, as it is trained on a compact set of informative markers. It operates much faster than most competing methods under study.

Sixth, as shown in real data applications, the proposed framework is scalable to large biobanks and genotype-imputed transcriptome data. It is designed to operate when only a subset of genes is available and when expression is derived from genetic prediction, enabling cell-type-resolved analyses at sample sizes that are typically infeasible for scRNA-seq.

Finally, a variety of downstream analyses are enabled by ***individual-level*** inference. Once cell-type proportions and CTS profiles are inferred per person under our framework, many analyses becomes possible, including CTS-TWAS, CTS direct causal gene detection via PC-simple, interaction models, prognosis modeling, and composition-adjusted TWAS.

### Related approaches

Numerous methods have been proposed to deconvolute cell compositions and estimate CTS expression profiles from bulk samples. However, these methodologies differ substantially from our current work. For instance, CIBERSORT/CIBERSORTx^9^ can infer cell compositions and CTS expression profiles from bulk transcriptomes. It utilizes reference gene expression profiles (GEPs) of predetermined marker genes for cell deconvolution. However, classical signature-matrix methods typically rely on the assumption of constant marker gene expression levels within cell types across samples, which might not hold in practice.

Another similar line of work is deep learning-based cell deconvolution and CTS expression estimation methods, such as Scaden and TAPE. Menden et al.^15^ introduced Scaden, a deep neural network-based cell deconvolution model designed to dissect bulk samples. Following a similar approach to Scaden, Chen et al.^16^ proposed an autoencoder-based model, TAPE, to infer cell compositions and CTS profiles from bulk samples. Rather than relying on GEPs of marker genes, they trained their deconvolution model directly on the reference scRNA-seq using typically ∼10,000 highly variable genes, making them computationally intensive and prone to learning dataset-specific artifacts or noise. CausalCellInfer occupies a strategic middle ground that combines the non-linear flexibility of deep neural networks with the rigorous, parsimonious feature selection of causal inference methods. Leveraging the invariance principle, the identified invariant marker genes from scI-GCM were likely more resistant to data bias and noises, and more generalizable to new datasets.

Another important distinction of our work from all previous studies on cell deconvolution is that we pioneered applications to genotype-imputed expression data, especially from large biobanks. Our work helps to bridge the gap between GWAS/sequencing and scRNA-seq studies and enables a wide variety of individual-level analysis with cell proportions and CTS expression and their relationship with diverse clinical phenotypes in a large sample. Such analyses are generally not possible in conventional single-cell studies due to the limited number of subjects and phenotypic information available.

More broadly, several frameworks link scRNA-seq and GWAS to identify disease-relevant cell types or to interpret GWAS signals in a cellular context. For example, Jagadeesh et al.^4^ integrated GWAS summary statistics, epigenomics and scRNA-seq data to detect the underlying cell types and processes by which genetic variants influence the target traits, and utilized stratified LD score regression to calculate heritability enrichment. In another study, scGWAS was developed to leverage scRNA-seq to interrogate CTS transcriptional changes induced by genetic variants^59^. It identified associations between traits and cell types by examining whether genes implicated by GWAS exhibit concordant activation in a particular cell type using proportion tests. Additionally, Zhou et al.^60^ proposed a CTS transcriptome-wide association framework (this work was released after our preprint was first posted in Oct 2024^61^). The approach leveraged external single-cell transcriptome and genotype data, and used Enformer-inferred epigenomics features to predict CTS expression; then a linearized model was built and association tests performed using GWAS summary statistics.

The current work is different from the aforementioned methods, which mainly operate with and focus on GWAS summary data. Our objective is fundamentally different: we infer ***individual-level*** cell-type proportions and CTS expression from bulk or genotype-imputed transcriptomes, enabling biobank-scale, cell-type-resolved association analyses across a wide phenotype spectrum. Individual-level data also allows for more diverse types of analysis, such as interaction modeling, cell-type-composition-adjusted TWAS, prioritization of potential direct causal genes via PC-simple, adjustment for other genetic/clinical covariates, flexible definition of outcome phenotypes, and the use of non-linear or machine learning models, which are generally not feasible with GWAS summary statistics alone. Moreover, our approach is methodologically novel and distinct, combining causal inference and invariance principles with deep neural networks and regularized matrix completion to estimate cell compositions and corresponding CTS expression profiles, a comprehensive strategy not previously employed.

### Limitations

Several limitations are worth noting but are likely increasingly addressable in practice. First, as with all reference-based methods, performance depends on the coverage and representativeness of the scRNA-seq reference. Rare cell types with few cells can yield less stable markers and less accurate cell fraction estimates. This challenge is expected to diminish as the volume of publicly available single-cell data grows, and it may be further mitigated by integrating multiple reference datasets and employing appropriate harmonization and batch correction methods. Furthermore, like most reference-based approaches, differences in factors such as ancestry, age, disease burden, and technical platforms between the reference and target cohorts (e.g., UK Biobank) can degrade transferability. Our proposed invariance-based marker selection aims to improve robustness and transferability across diverse environments, though it cannot entirely eliminate the effects of such mismatches.

Second, genotype-imputed expression reflects only the genetically regulated component of expression, and typically has reduced dynamic range relative to measured bulk RNA-seq. Our pseudo-bulk calibration provides a pragmatic way to align scales for deconvolution and CTS reconstruction, but future work could further refine this step or incorporate uncertainty in imputation.

Third, similar to the principles of Mendelian Randomization (MR), our analysis is based on genetically predicted expression, and focuses on how expression influences outcomes. The approach does not capture reverse causality, where phenotypic traits might induce changes in gene expression. Also, our characterization of identified genes and markers as “causal” is contingent upon satisfying the standard assumptions of causal graphical models and invariant prediction. Consequently, while our findings are promising, these outputs should be interpreted as hypothesis-generating and putative, requiring experimental validation for definitive confirmation. It is worth noting, however, that markers identified as invariant are expected to be less sensitive to context-induced shifts. Therefore, even if they are not strictly causal, they are likely to be more transferable and robust across different cohorts and conditions. Despite the above limitations, our proposed framework is highly flexible and holds potential for extension to address these challenges in future developments.

### Conclusions

In summary, CausalCellInfer offers a scalable strategy for converting bulk and genotype-imputed transcriptomes into *cell-type-resolved* readouts. This enables a wide variety of analyses at biobank scale, such as systematic CTS-TWAS, cell type composition-aware inference, interaction analyses, and causal gene prioritization. Through benchmarking and application to the UK Biobank, we demonstrate that cellular hypotheses can be tested with rich phenotypic depth, facilitating the translation of genomic associations into mechanistic and clinically actionable insights.

## Data availability

UK biobank data is available to any researchers who formally apply for the data. However, the data is not publicly available due to privacy concerns. Reference scRNA-seq datasets are publicly available at the following sites: frontal cortex: https://www.ncbi.nlm.nih.gov/geo/query/acc.cgi?acc=GSE144136 adipose tissues: https://singlecell.broadinstitute.org/single_cell/study/SCP1376/a-single-cell-atlas-of-human-and-mouse-white-adipose-tissue?cluster=Human%20WAT&spatialGroups=--&annotation=fat_type--group--study&subsample=100000#study-summary pancreas: https://hpap.pmacs.upenn.edu/

## Code availability

The source codes and R package to reproduce our experiments for this work is available via GitHub: https://github.com/yujias424/CausalCellInfer.

## Conflicts of interest

The authors declare no relevant conflicts of interest.

## Acknowledgements

This work was supported partially by a Young Collaborative Research Grant (C4003-23Y), Lo Kwee Seong Biomedical Research Fund from The Chinese University of Hong Kong and the KIZ-CUHK Joint Laboratory of Bioresources and Molecular Research of Common Diseases, Kunming Institute of Zoology and The Chinese University of Hong Kong, China. An earlier version of this manuscript was released as a preprint on medRxiv on 17 Oct 2024. (https://doi.org/10.1101/2024.10.17.24315646)

## Notes

### Competing Interest Statement

The authors have declared no competing interest.

### Funding Statement

This work was supported partially by the Lo Kwee Seong Biomedical Research Fund from The Chinese University of Hong Kong and the KIZ-CUHK Joint Laboratory of Bioresources and Molecular Research of Common Diseases, Kunming Institute of Zoology and The Chinese University of Hong Kong, China.

### Author Declarations

North West - Haydock Research Ethics Committee gave ethical approval for this work

### Summary of Updates

I have updated the introduction, methods, and results sections. Some content in the method section has been moved to the supplementary text. I have deleted one supplementary table.

