## Supplementary Text for "CausalCellInfer: Resolving cell-type-specific disease mechanisms from biobank-scale GWAS"

#### UK Biobank data and phenotype definitions

UKBB is a comprehensive database that provides extensive prognostic information on various diseases. Using cell-type-specific (CTS) expression profiles, we investigated the relationship between cell-specific genes and disease prognoses, including mortality and hospitalization frequency. Our application to UKBB focused on 4 primary tissue-specific diseases/outcomes (i.e., major depression in the frontal cortex, T2DM in the pancreas, and obesity in subcutaneous and visceral adipose tissues). We also studied 8 prognosis-related variables and 17 related disorders or traits. Here we concentrated on prognosis variables that are of high clinical significance for the target diseases.

More specifically, we focused on treatment resistance status and depression-related hospitalization frequency for patients with depression (tissue studied: frontal cortex); T2DM-related mortality and hospitalization frequency (tissue studied: pancreas) in T2DM subjects; coronary artery disease (CAD) mortality, and CAD-related hospitalization frequency for obesity patients (tissue studied: subcutaneous and visceral adipose tissues[SAT/VAT]). We also studied other related disorders, including anxiety disorders, bipolar disorder, and psychosis (frontal cortex); prediabetes, T1DM, T2DM requires hospitalization, T2DM requires insulin (pancreas); CAD, T2DM, hypertension, heart failure, stroke (SAT/VAT) respectively.

With the UKBB summary data of hospital inpatient episodes (Data-Field 41270) and their corresponding diagnosis dates (Data-Field 41280), we extracted relevant phenotypes based on the International Classification of Disease version 10 (ICD-10) code. For each disorder, cases were defined as participants with either an ICD-10 coded primary diagnose or a corresponding self-reported diagnosis; controls were participants without any record of that code or related self-report.

##### *Depression, Depression related disorders and prognosis variables*

Depression and related disorders were directly extracted from the UKBB with a combination of self-reported data and ICD-10 codes. Depression is characterized by ICD-10 codes F32-33; psychosis is characterized by ICD-10 codes F20-25; bipolar disorder is characterized by F31; anxiety disorders are characterized by ICD-10 code F41.

Using the general practitioner (GP) records, we also extracted treatment-resistance status and admission frequency of depression patients. Specifically, treatment-resistant depression (TRD) was defined as depression patients who tried at least two different anti-depressant drugs for adequate durations. Following the definition in[1], the time interval between two drugs should be no longer than 14 weeks and each drug should be prescribed for at least 6 weeks.

***T2DM, T2DM-related disorders and prognosis variables***

The prediabetes phenotype is specifically characterized by codes starting with R73, whereas the ICD-10 codes for T1DM and T2DM begin with E10 and E11 respectively. For each patient with available hospital records, the earliest diagnosis dates for prediabetes, T1DM, and T2DM were extracted for data analysis. T2DM-related admission was defined as T2DM patients who had at least one hospitalization record related to T2DM.

Besides, by restricting the Anatomical Therapeutic Chemical (ATC) code starting with A10A, we obtained the issue date of insulin from General Practice (GP) records, which contain prescription data from two electronic health record (EHR) systems (TPP or EMIS) for UKBB participants. In our “T2DM requiring insulin” dataset, once insulin is prescribed after the earliest T2DM diagnosis, the variable “T2DM requiring insulin” was coded as 1, otherwise it was coded as 0.

***Obesity, obesity related disorders and prognosis variables***

For obesity, we adhered to the World Health Organization (WHO) definition, which classifies individuals with a Body Mass Index (BMI) exceeding 30 as obese. CAD, hypertension, stroke, heart failure and T2DM were directly extracted from the UKBB with a combination of self-reported data and ICD-10 codes.

**Methods**

***I-GCM***

Briefly, I-GCM is a two-stage approach, which firstly conducts feature selection using PC-simple, then leverages the principle of causal invariance across different environments to identify robust direct causal variables. Briefly, PC-simple can be regarded as a generalization of ordered-correlation screening, which is used to preselect the direct causal variables. Suppose $X = [X^1, X^2, \dots, X^p]$ is a $n \times p$ matrix for $n$ observations with $p$ variables, $Y$ is the target variable. Variable $X^j$ is independent of Y if:

$$\exists s \subseteq S, \rho(Y, X^j|X^s) = 0 \tag{1}$$

$S = \{1,2, \dots, j-1, j+1, \dots, p\}$ defines the set including variables excluding $X^j$, $s$ is a subset of $S$, $\rho(Y, X^j|X^S)$ denotes the partial correlation between $X^j$ and $Y$. The null hypothesis of independence would be rejected if

$$(n - |s| - 3)^{1/2}|Z(Y, X^j|X^S)| > \phi^{-1}(1 - \alpha/2) \tag{2}$$

Where $Z(Y, X^j|X^s) = \frac{1}{2}\{\frac{1+\hat{\rho}(Y, X^j|X^s)}{1-\hat{\rho}(Y, X^j|X^s)}\}$ denotes Fisher's Z-transform of the partial correlation, $n$ denotes the sample size, $|s|$ is the cardinality of the set, $\phi$ indicates the inverse cumulative function for normal distribution. In our implementation, $Y$ denotes the cell-type indicator, $X$ indicates the gene expression matrix. Recursively performing partial correlation screening with increased order could exclude irrelevant variables from previous candidate variables set until they do not vary anymore. In this study, we set the maximum order of $|s|$ to 3 and $\alpha$ to 0.05 for the feature screening process. This implies that variables which survive the 3-order partial correlation screening will be retained for further analysis. The corresponding p-value of the independence test can be used to indicate the reliability of the identified causal variable. The smaller the p-value, the more reliable the identified causal relationship is. As have been proven by Rajen et al.[1], generalized covariance measure (GCM) (a.k.a., $T^n$) can also be used to test whether $X^j$ and Y are conditional independent given variable(s) $X^S$ :

$$T^n = \frac{\sqrt{n}.\frac{1}{n}\sum_{i=1}^{n} R_i}{(\frac{1}{n}\sum_{i=1}^{n} R_i^2 - (\frac{1}{n}\sum_{r=1}^{n} R_r)^2)^{1/2}} = \frac{\frac{1}{n}\sum_{i=1}^{n} R_i}{[(\frac{1}{n}\sum_{i=1}^{n} R_i^2 - (\frac{1}{n}\sum_{r=1}^{n} R_r)^2)^{1/2}]/\sqrt{n}} \tag{3}$$

Where $R_i = (X_i^j - f(X_i^S))(Y_i - f(X_i^S))$ defines the product between residuals($R$) from the prediction functions of $X^j$ and Y using $X^S$ for each observation. In this study, XGBoost was employed to build prediction models. XGBoost[2], a supervised machine learning method, is composed of a sequence of simpler, weaker decision trees. It operates by iteratively adding new trees, each one designed to correct the errors made by its predecessors, until the model achieves an optimal fit to the data. The complexity of the trees is penalized to prevent overfitting and enhance the model's generalizability.

For a given environment variable E(i.e., disease status, sex), the computed GCM between $E$ and the target should be constant or similarly close to zero when conditioned on the full set of direct causes, even if some other (non-directly causal) variables are included. However, if we exclude some direct causes from the conditional set, the calculated GCM between $E$ and the target is expected to divert away from the null. In other words, significant change of the calculated GCM would be observed between the reduced conditional set and full conditional set. If we exclude some irrelevant variables in the conditional set, the calculated GCM should remain stable. While disease status was selected as the environment variable for pancreas, frontal cortex, subcutaneous adipose tissue, visceral adipose tissue, batch was selected for peripheral blood mononuclear cells tissue. The change in the distance of GCM from zero between two consecutive conditional sets ($\Delta T^n_{S_{j-1},S_j}$) could be represented as follows:

$$\Delta T^n_{S_{j-1},S_j} = |T^n_{S_{j-1}}| - |T^n_{S_j}| \tag{4}$$

In this study, $\alpha$ was set to 0.05. The variance of the distance change of GCM (Var($\Delta T^n_{S_{j-1},S_j}$)) under the null can be estimated from the following:

$$\begin{aligned}\mathrm{Var}(\Delta T^n_{S_{j-1},S_j}) &= Var\left(\left|\left|T^n_{S_{j-1}}\right|\right|\right) + Var\left(\left|T^n_{S_j}\right|\right) - 2\left[E\left(\left|T^n_{S_{j-1}} * T^n_{S_j}\right|\right) - E\left(\left|T^n_{S_{j-1}}\right|\right) * E\left(\left|T^n_{S_j}\right|\right)\right] \\ &= 2 - \frac{4}{\pi}\left(\rho * arcsin\rho + \sqrt{1-\rho^2}\right) \text{ (see refs[3,4])}\end{aligned} \tag{5}$$

where $\rho = Cor(T^n_{S_{j-1}}, T^n_{S_j})$ indicates the correlation between product of residuals calculated from conditional set $S_{j-1}$ and $S_j$, $S_{j-1}$ and $S_j$ respectively denote two consecutive conditional set with $j-1$ and $j$ variables.

The null hypothesis of no significant distance change in GCM between two consecutive conditional sets would be rejected if:

$$\frac{\Delta T^n_{S_{j-1},S_j}}{sqrt\left(\mathrm{Var}\left(\Delta T^n_{S_{j-1},S_j}\right)\right)} > \phi^{-1}(1-\alpha) \tag{6}$$

#### *scI-GCM*

In this study, we proposed to employ a modified I-GCM method designed for single cell data (scI-GCM) to identify cell markers. Algorithm 1 summarizes how we identify the corresponding cell markers for each target cell type using scI-GCM.

**Algorithm S1: scI-GCM for cell marker identification**

**Input**: $\boldsymbol{X} \in \boldsymbol{R}^{n \times q}$, $\boldsymbol{Y} \in \boldsymbol{R}^{n}$ (cell type labels), $E \in \boldsymbol{R}^{n}$, $\boldsymbol{\alpha}$, $T$

**For each cell type** $\boldsymbol{m} = \boldsymbol{c_1} : \boldsymbol{c_m}$

  *If* $\boldsymbol{Y_i} = \boldsymbol{m}$ set $\boldsymbol{Y_i} = \boldsymbol{1}$ otherwise set $\boldsymbol{Y_i} = \boldsymbol{0}$

  For each environment $\boldsymbol{e}$:

    Identify candidate marker genes using XGBoost(environment variable e excluded)

    Further remove redundant marker genes using PC-simple to get marker gene set for environment $\boldsymbol{e}$: $\boldsymbol{G_e}$

  **end** Merge and re-rank $\boldsymbol{G_e}$ based on the derived Z-score from PC-simple in descending order to get the candidate marker gene set: $\boldsymbol{G_q}$

  For j = **q**: **2**, set $\boldsymbol{S_j} = \{\boldsymbol{1}, \boldsymbol{2}, \ldots \boldsymbol{j}\}$, $\boldsymbol{S_{j-1}} = \{\boldsymbol{1}, \boldsymbol{2}, \ldots \boldsymbol{j} - \boldsymbol{1}\}$

1. Calculate the GCM between $\boldsymbol{E}$ and $\boldsymbol{Y}$ by conditioning on $\boldsymbol{S_j}$ ($\boldsymbol{T^n_{S_j}}$) and $\boldsymbol{S_{j-1}}$($\boldsymbol{T^n_{S_{j-1}}}$) respectively based on equations 3
2. Compute the distance change between $\boldsymbol{T^n_{S_{j-1}}}$ ***and*** $\boldsymbol{T^n_{S_j}}$, and the corresponding variance based on equations 4-5
3. Test the null hypothesis of no significant distance change. If inequality holds(inequality 6) and $\Delta \boldsymbol{T^n} > \boldsymbol{T}$ stop the testing

**end**

**Output**: marker gene set for cell type $\boldsymbol{m}$ : $\boldsymbol{S_m}$

$\boldsymbol{X} \in \boldsymbol{R}^{n \times q}$ : gene expression matrix with n indicates cell number, q indicates gene number; $\boldsymbol{Y} \in \boldsymbol{R}^{n}$: cell type labels; $E \in \boldsymbol{R}^{n}$: environment variable; $\boldsymbol{m}$ is the number of total cell types; $\boldsymbol{S_m}$: identified cell markers for the corresponding cell type $m$; after ranking, indices $1, \ldots, q$ correspond to genes from most to least strongly associated. $\boldsymbol{S_j}$ thus denotes the top j genes in this ranked list; $T$ is set to 0 for this task.

### Evaluation of the proposed scI-GCM approach

To evaluate the performance of the scI-GCM approach in handling scRNA-seq datasets confounded by environmental variables, we designed a simulation experiment to deconvolute a series of synthetic bulk samples comprising four cell types with varying confounding effect sizes. We utilized scDesign3[2] to generate synthetic scRNA-seq data based on a publicly available peripheral blood mononuclear cell (PBMC) dataset containing known batch effects. To reduce computational cost, we selected four cell types from the PBMC dataset and restricted the simulation to 200 genes. Using the reference PBMC data, we fitted an interpretable parametric model in scDesign3, capturing both marginal distributional parameters and pairwise gene correlations. A key advantage of scDesign3 is its flexibility; it allows for the modification of fitted parameters to reflect specific hypotheses and generate corresponding in silico data that retain real-world characteristics. Leveraging this, we generated a baseline scRNA-seq dataset free of batch effects by setting the batch covariate coefficients to zero. Subsequently, we generated datasets with batch effects by adjusting these coefficients to specific constants. We simulated two scenarios representing moderate and strong batch effects by setting the coefficient magnitudes to 8 and 16, respectively. Finally, following the TAPE approach, we simulated bulk samples derived from both the batch-free and batch-affected scRNA-seq datasets. Comprehensive simulation code and scripts are available in the accompanying GitHub repository.

### PC-simple algorithm

We employed PC-simple to detect cell-type specific genes that are likely directly causal to the studied phenotypes. PC-simple is capable of distinguishing direct causal variables from indirect causal and spuriously associated genes. It can be considered a localized version of the PC algorithm, where an outcome (*Y*) and a predictor are deemed causally linked if they remain correlated, even when conditioned on any subset of the other covariates.

Suppose *Z* and *Y* are two random variables, and *S* is a variable set excluding *Z* and *Y*. *Z* and *Y* are conditionally independent if:

$$\exists s \subseteq S, \rho(Z, Y|S) = 0$$

$\rho(Z, Y|S)$ denotes the partial correlation between $Z$ and $Y$ given S. Let $X = [X^1, X^2, \dots X^p]$ be a $n \times p$ matrix of adjusted gene expression data for $p$ genes, *Y* be a vector of the corresponding adjusted phenotypes for *n* subjects. We could identify the gene sets that are strongly associated with the target $Y$ given other variables at a specified significance level $\alpha$ by ordered independence screening.

Initially, the candidate set includes all input variables. We iteratively update the candidate set by removing independent variables via conditional independence tests. The conditional independence tests are done level by level with an increasing order of the conditional set, starting with an empty conditional set. As shown in Algorithm S1, the initial candidate set $\boldsymbol{G^0}$ is the complete set of input variables. Candidate set $\boldsymbol{G^1}$ is firstly set to $\boldsymbol{G^0}$ and then updated by removing independent variables via independence tests (conditioned on the null set). Similarly, candidate set $\boldsymbol{G^2}$ is initially set to $\boldsymbol{G^1}$, and is updated by removing variables that are independent of $Y$ given any other single variable in $\boldsymbol{G^1}$. Through recursively performing independence screening with an increasing order of the conditional set, we exclude genes from the previous candidate gene set until it does not change anymore.

To improve computational efficiency, we set the maximum order for partial correlation screening to 3. Fisher's Z-transformation was employed to convert the skewed partial correlation into a normally distributed variable, and we tested whether the partial correlation is significantly different from zero:

$$Z_{score}(Y, Z|S) = \frac{1}{2}\{\frac{1 + \hat{\rho}(Y, Z|S)}{1 - \hat{\rho}(Y, Z|S)}\}$$

As suggested by Buhlmann et al. [3], the null hypothesis $\hat{\rho}(Y, Z|S) = 0$ would be rejected if

$$(n - |S| - 3)^{1/2} |Zscore(Y, Z|S)| > \phi^{-1}(1 - \alpha/2)$$

where $|S|$ defines the order of the conditional set, $\alpha$ and $\phi$ respectively denote the significance level and standard normal cumulative distribution function. In this study, we set $\alpha$=0.05. The derived vector $Z_{p\times1}$ indicates the reliability of the inferred causal relationships between variables. We could convert it into a $p$-value vector by $P = 2 * \phi(-abs(Z))$.

**Calibrating PrediXcan-imputed expression**

The scale of the PrediXcan data is incompatible with scRNA-seq reference data, preventing direct inference of cell composition and cell-type-specific expression.

To mitigate this limitation, we mapped the PrediXcan-imputed values back into the expression space of the scRNA-seq reference, using a heuristic pseudo-bulk calibration. Specifically, we generated 1,000 pseudo-bulk RNA-seq samples from the control-group scRNA-seq data using a procedure similar to that used in the TAPE approach[4]:

1. We took the reference cell-type proportions as a baseline. For each pseudo-bulk sample, we perturbed these proportions by adding Gaussian noise to each cell type, clipped negatives to zero, and renormalized to sum to one.
2. We then sampled 1,000 cells from the scRNA-seq reference with probabilities given by the perturbed proportions and summed their profiles to form one pseudo-bulk sample.

From these 1,000 pseudo-bulk samples, we computed, for each gene $g$, the mean $\mu_g$ and standard deviation $\sigma_g$ . For each subject $s$, let $Z_{g,s}$ denote the PrediXcan-imputed standardized value for gene $g$. We define the calibrated pseudo-bulk-like value:

$$\tilde{O}_{g,s} = Z_{g,s}\,\sigma_g + \mu_g.$$

We use $\tilde{O}$ as the bulk input $O$ for both DNN deconvolution and ENIGMA reconstruction.

This approach relies on two key assumptions. First, we assume that standardized PrediXcan imputations, trained on normal-tissue RNA-seq data (e.g., GTEx), accurately reflect genetically regulated expression and preserve the hierarchy of gene expression across individuals within a tissue. Second, we assume that the cell proportion ratios and cell-type-specific expression profiles derived from reference single-cell RNA-seq (scRNA-seq) data of healthy individuals follow a distribution similar to that of the normal-tissue RNA-seq data used for PrediXcan training. Under these assumptions, mapping PrediXcan-imputed values onto a pseudo-bulk scale estimated from control scRNA-seq data allows for the recovery of both between-individual variation and within-individual gene expression rankings. For instance, housekeeping genes remain highly expressed, while lowly expressed genes remain relatively silent; similarly, genes highly expressed in specific population subgroups retain their elevated status after mapping. The resulting rescaled bulk RNA-seq data are subsequently used for cell type proportion estimation and the inference of cell-type-specific expression profiles.

We note that this mapping procedure serves as a heuristic calibration step: it posits that the standardized PrediXcan-predicted values for each gene exist on a latent scale comparable to the pseudo-bulk expression distribution estimated from the scRNA-seq reference. Nevertheless, this offers a practical means to generate bulk-like expression magnitudes compatible with ENIGMA or other cell deconvolution approaches, all while preserving the population-wide gene expression hierarchy described above.
